# Farm-to-consumer quantitative microbial risk assessment model for *Listeria monocytogenes* on fresh-cut cantaloupe

**DOI:** 10.1101/2025.03.01.25323161

**Authors:** Sarah Ingersoll Murphy, Ece Bulut, Laura K. Strawn, Michelle D. Danyluk, Martin Wiedmann, Renata Ivanek

## Abstract

Cantaloupe contamination with the foodborne pathogen *Listeria monocytogenes* (LM) may occur along the supply chain. We developed a quantitative microbial risk assessment (QMRA) model for LM on cantaloupe along the fresh-cut supply chain and evaluated potential risk reduction strategies. The developed model starts at harvest and includes conditions during transportation from field to intermediate facility (packinghouse or cooling facility), handling at the intermediate facility, transportation to the fresh-cut facility, storage pre-processing at the fresh-cut facility, processing and handling at the fresh-cut facility, as well as conditions during distribution, retail, transportation to home, and home storage. The model was simulated to (i) provide an estimate of LM concentration in a single serving (134 g) and (ii) estimate annual illnesses and deaths in the United States attributed to LM contaminated fresh-cut cantaloupe. The baseline model predicted the median risk of listeriosis per serving in general and susceptible populations was 1.4 × 10^−12^ and 6.4 × 10^−11^, respectively. The median (5^th^, 95^th^ percentiles) predicted number of illnesses and deaths annually attributed to fresh-cut cantaloupe were 0 (0, 1070) and 0 (0, 264), respectively. Time and temperature conditions post-packaging, and the initial number of LM at harvest had the greatest impacts on LM per contaminated serving and the number of annual illnesses; the initial LM levels at harvest and cross-contamination parameters at the fresh-cut facility had the greatest impacts on prevalence of contaminated servings. Assessment of interventions demonstrated that reducing temperature and/or time conditions post-packaging can be an effective risk reduction strategy. Overall, the developed tool estimates the risk associated with the consumption of LM contaminated fresh-cut cantaloupe and facilitates the identification and assessment of potential risk reduction strategies across the supply chain.

## INTRODUCTION

*Listeria monocytogenes* (LM) is a foodborne pathogen that is of increasing concern for the produce industry, due to human health risk as well as potential for negative impacts on the industry. For example, after a 2011 United States (US) listeriosis outbreak, with 147 illnesses and 33 deaths linked to consumption of contaminated cantaloupe originating from a single farm (*21*), US cantaloupe consumption dropped by 53% (*1*). Similarly, a 2018 Australian listeriosis outbreak attributed to cantaloupe from a single farm that resulted in 22 illnesses and 7 deaths (*25*) had negative impacts on the industry overall (e.g., loss of sales) (*3*). In response to the identified risk of LM on cantaloupe, guidance has been developed and provided to industry for food safety management from pre-harvest through retail (*9, 31, 40*). Notably, the majority (21/28) of cantaloupe-related LM outbreaks reported to CDC from 1973-2003 were linked to fresh-cut cantaloupe (*21*). While the previously recorded LM outbreaks have been linked to whole cantaloupe, there is additional food safety risk for fresh-cut relative to whole cantaloupe, for example due to the additional handling.

LM has been previously detected in each stage of the fresh produce supply chain (*37*). Cantaloupe is susceptible to contamination with LM at various points across the supply chain, such as from sources in the natural environment at the field-level or interactions with contaminated workers or equipment during processing and handling. Furthermore, studies have shown LM is able to grow on both cantaloupe rind and flesh (i.e., whole cantaloupe and fresh-cut cantaloupe) (*7, 17, 30*), highlighting a need for risk reduction strategies throughout the supply chain (e.g., temperature controls). While many postharvest control strategies have been suggested, their relative efficacy is not well understood, thus making it difficult to select appropriate strategies to implement.

Previous studies have developed quantitative risk assessments to inform risk management of cantaloupe, demonstrating the utility of this approach (*6, 22*). For example, Mokhtari et al. (*22*) developed a quantitative microbial risk assessment (QMRA) of human health risk from *Salmonella* on cantaloupe from the fresh-cut facility through storage and consumption at home, presenting the QMRA as a tool for assessment of mitigation strategies at the fresh-cut facility level. In their report presenting US Food and Drug Administration (FDA)’s modeling tool for food safety risk assessment, FDA-iRisk, Chen et al. (*6*) presented a set of case studies for LM and *Salmonella*, including a risk assessment for LM on cantaloupe for US adults over 60 years old that only included home storage. While these studies provide valuable insights into the pathogen risks at various stages of the supply chain and across different populations, a QMRA that covers the entire farm-to-consumption pathway for LM on cantaloupe for the general and susceptible population is still lacking. To address this gap, the goal of the study reported here is to develop a QMRA of human health risk from LM on cantaloupe along the fresh-cut supply chain, from farm-to-consumption, to facilitate a systems-approach for identification of appropriate postharvest risk reduction strategies. While cantaloupe handling practices vary in the US, our study focuses on two supply chain scenarios: (i) transportation to a packinghouse where cantaloupes are washed, repacked, and then sent to a fresh-cut facility for washing, cutting, and packaging for retail distribution, and (ii) “field packing” followed by transportation to a cooling facility for storage before being transferred to a fresh-cut facility. Overall, the developed QMRA will allow for estimation of illnesses and deaths associated with the consumption of LM-contaminated fresh-cut cantaloupe in the US and will facilitate identification and evaluation of the relative impact of risk reduction strategies.

## MATERIALS AND METHODS

### Model overview

A conceptual map showing the steps and basic processes affecting prevalence and concentration is provided (Figure 1). A summary of all model variables, baseline parameter values, and their sources are provided in Tables 1 and 2. The model starts with a random ½ ton bin of harvested cantaloupes at the field-level, with an initial probability of a bin containing LM-contaminated cantaloupe (*P*_0_,_*Bin*_) and initial number of LM cells on the total cantaloupe surface area in a contaminated bin (*N*_0_,_*Bin*_). Due to lack of available data no other steps were modeled at the field-level. We assumed that cantaloupes were packed at the field and then transported either to a cooling facility (Scheme A) or a packinghouse (Scheme B). For Scheme B, at the packinghouse, cantaloupes were washed and then repacked (no washing and repacking at Scheme A), which was simulated as inactivation and cross-contamination in the packinghouse environment. Both Scheme A and B included storage. Cantaloupes were then transported from the intermediate facility (i.e., cooling facility or packinghouse) to the fresh-cut facility and then stored until processing. At the fresh-cut facility cantaloupes were washed, cut, and packed, which was simulated as inactivation, cross-contamination in the processing environment, partitioning (recognizing individual cantaloupes in the bin), transfer (rind to flesh), and partitioning (cantaloupes to packages). Fresh-cut cantaloupe packages were then distributed (transportation and storage) to retail stores (storage and display), followed by transportation from retail to home and then home storage. Finally, an individual serving of fresh-cut cantaloupe was prepared from package, which was modeled as partitioning.

**Table 1.**
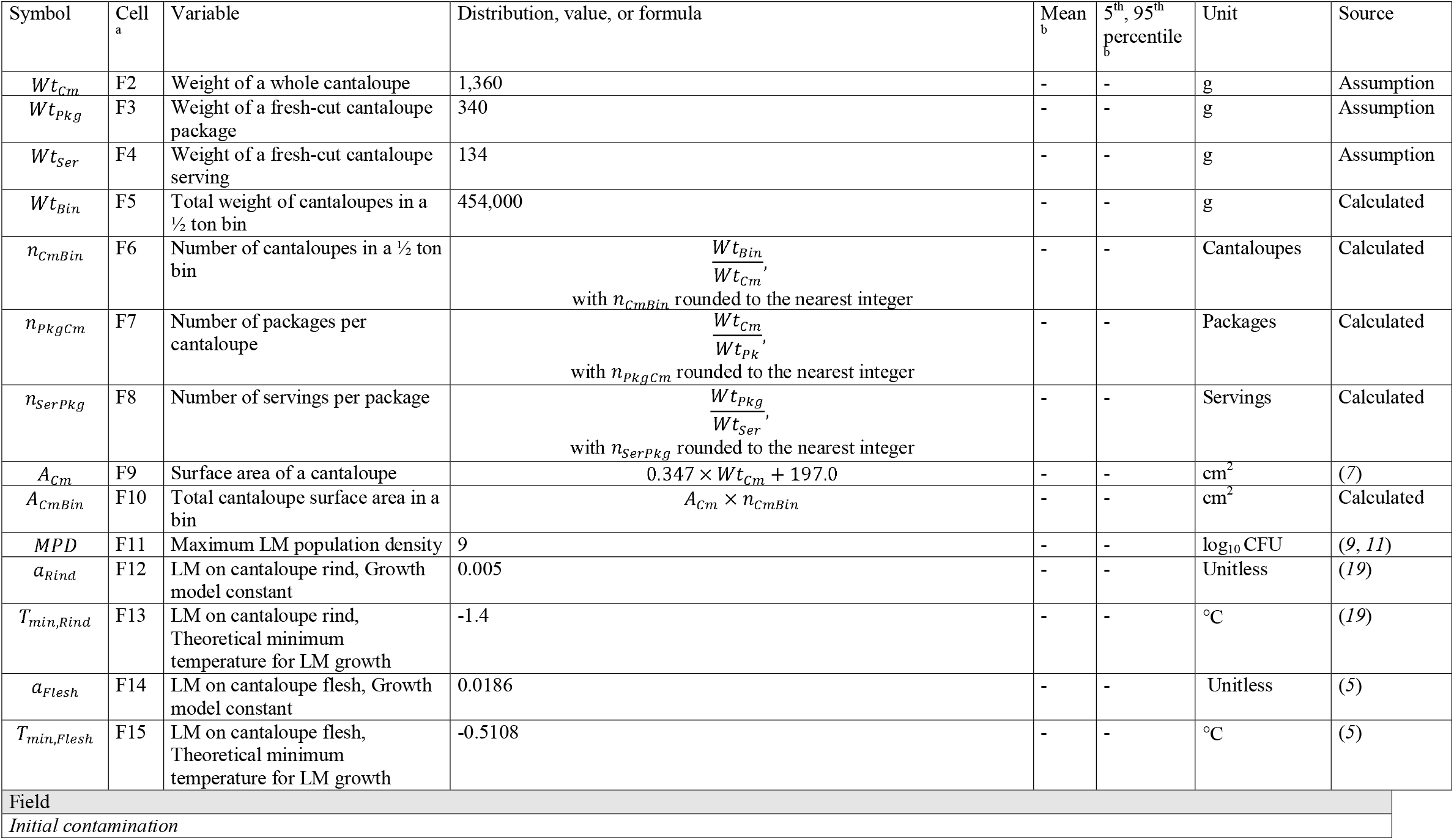
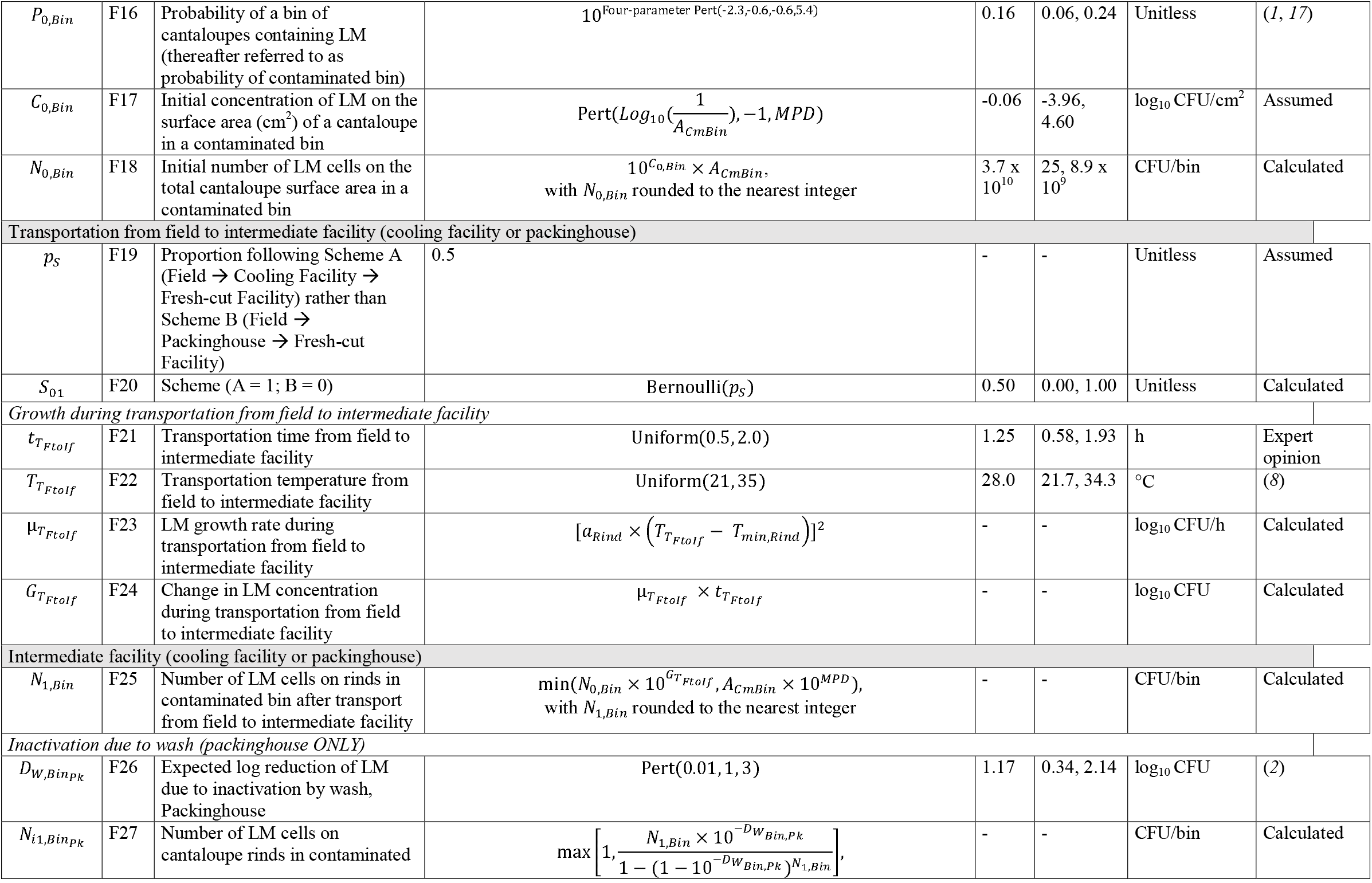
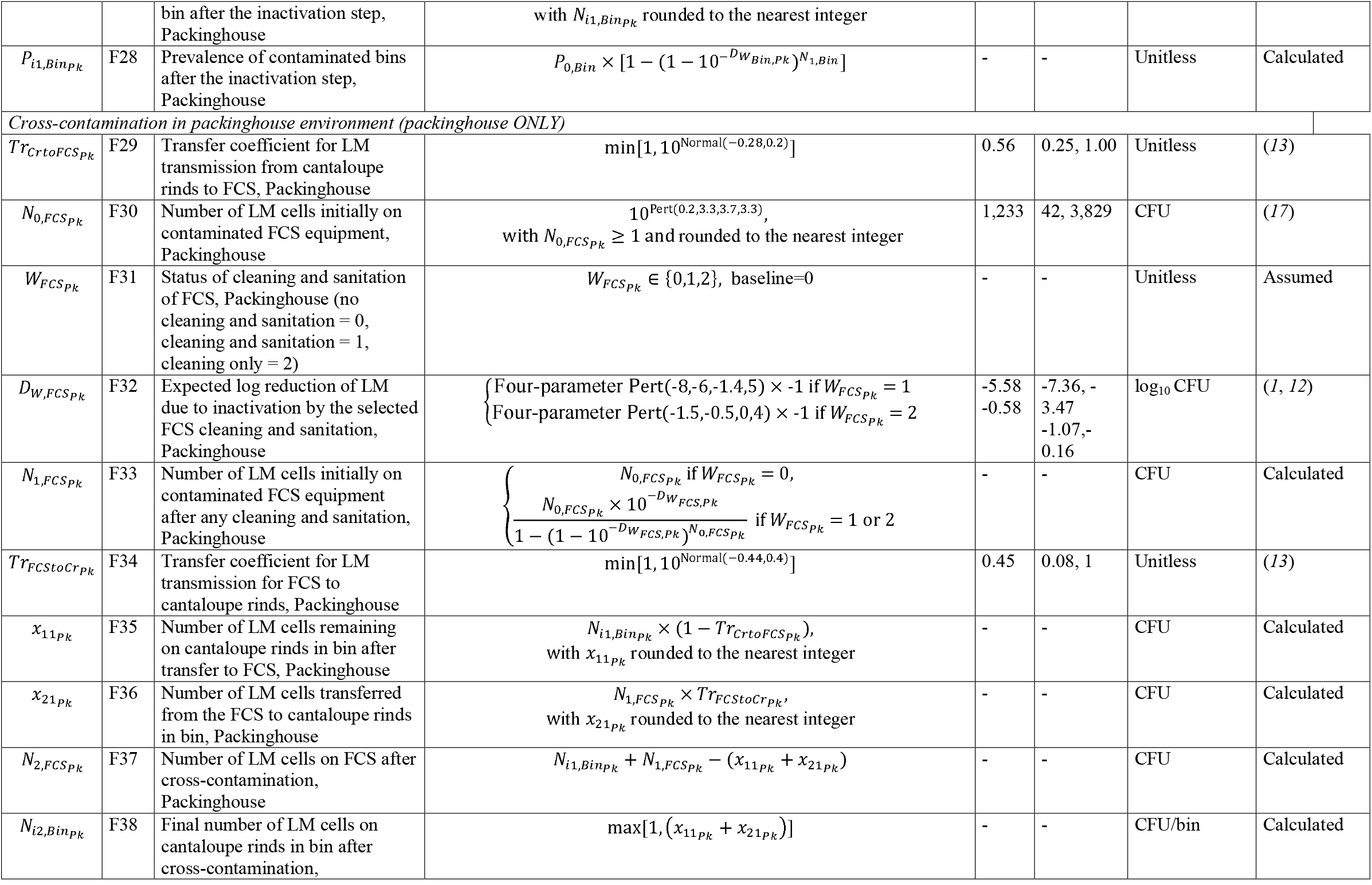
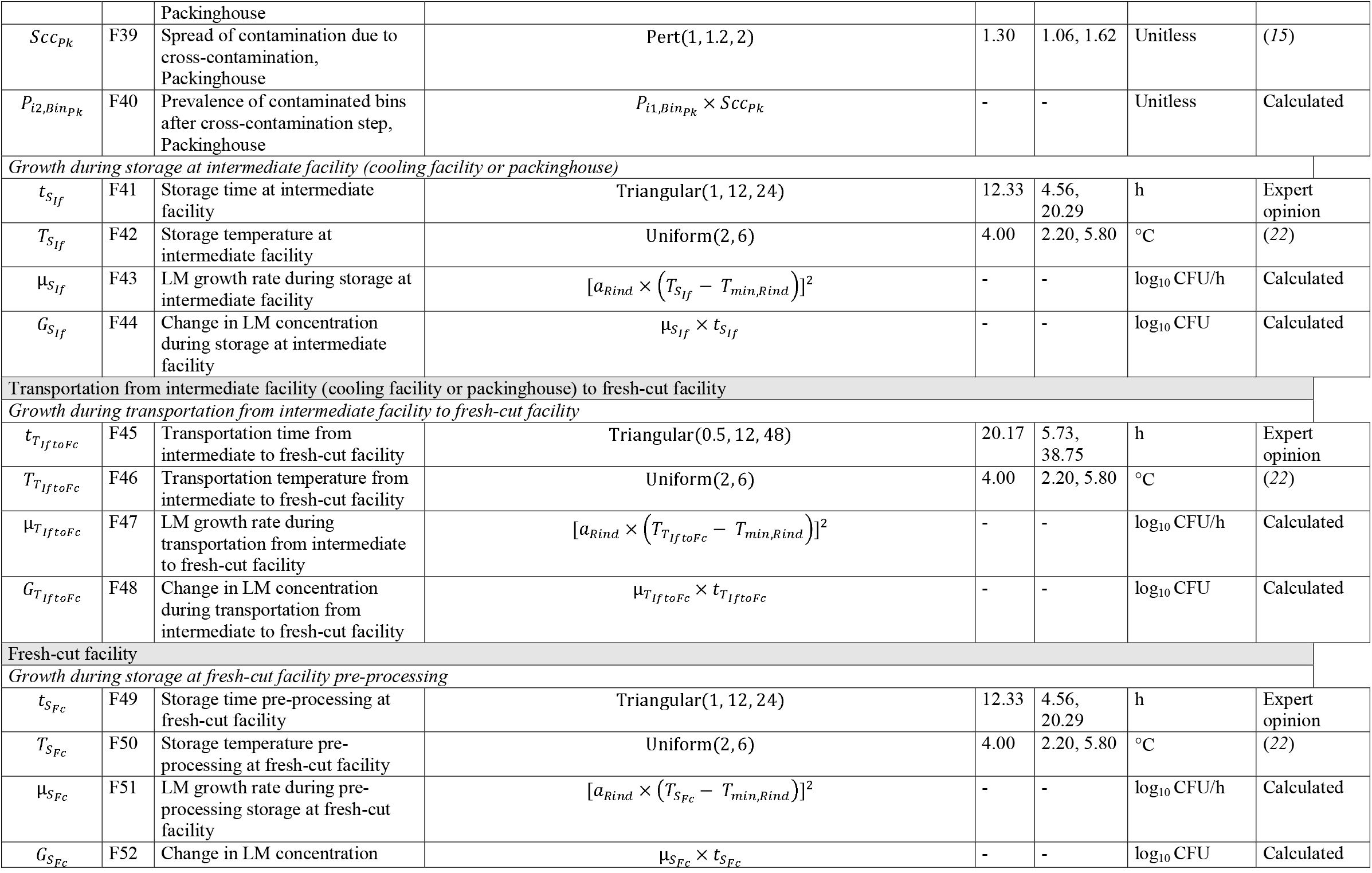
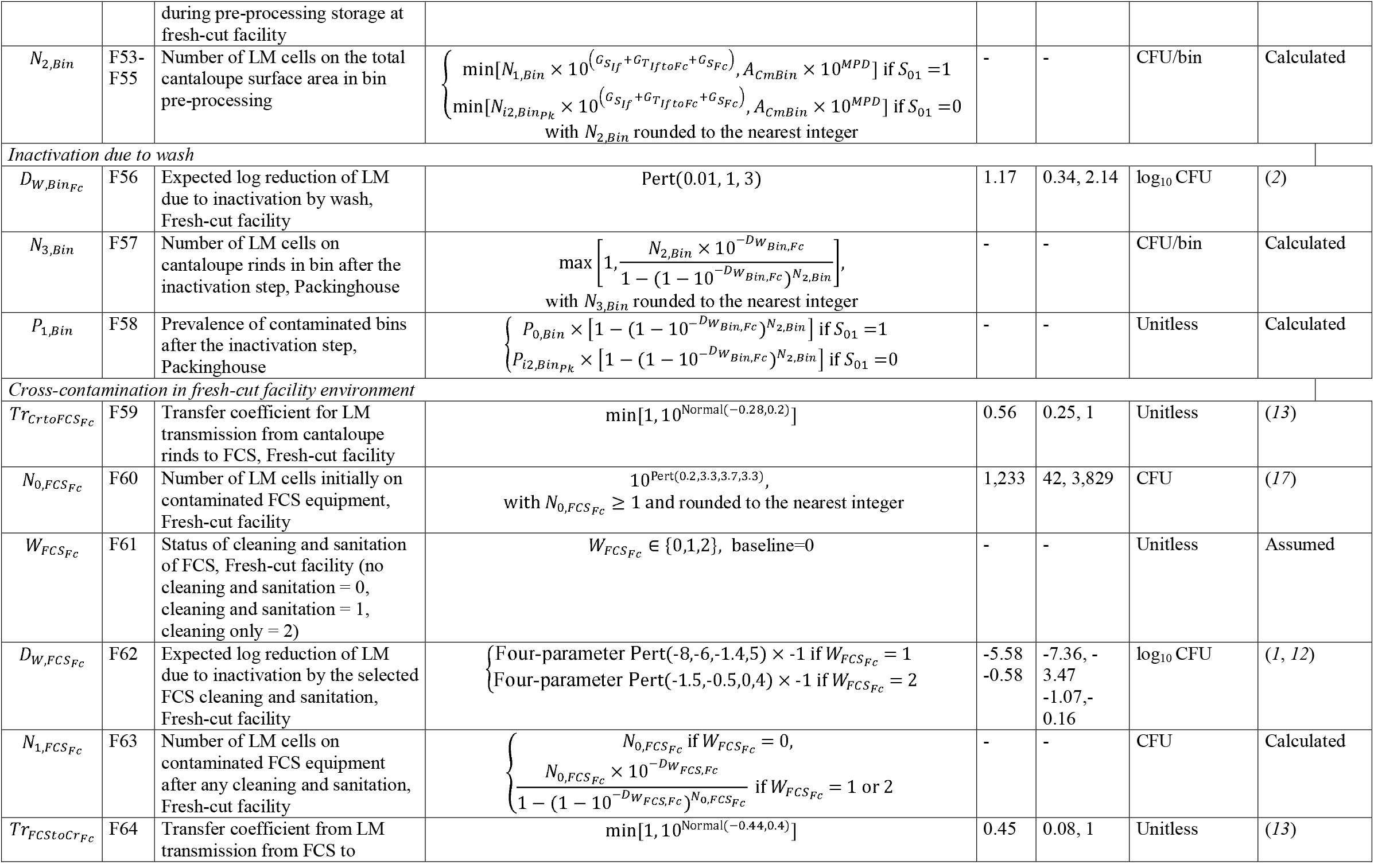
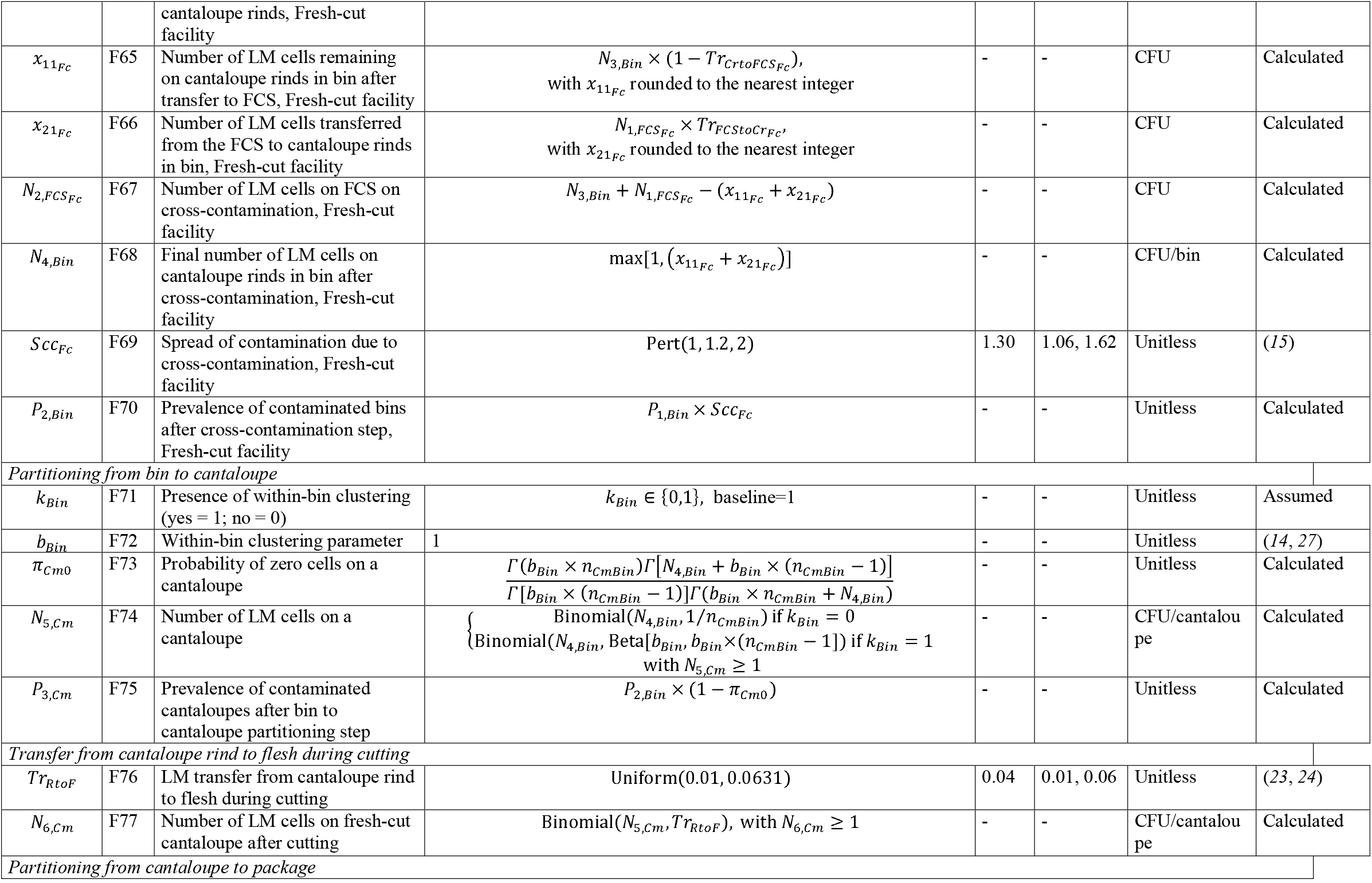
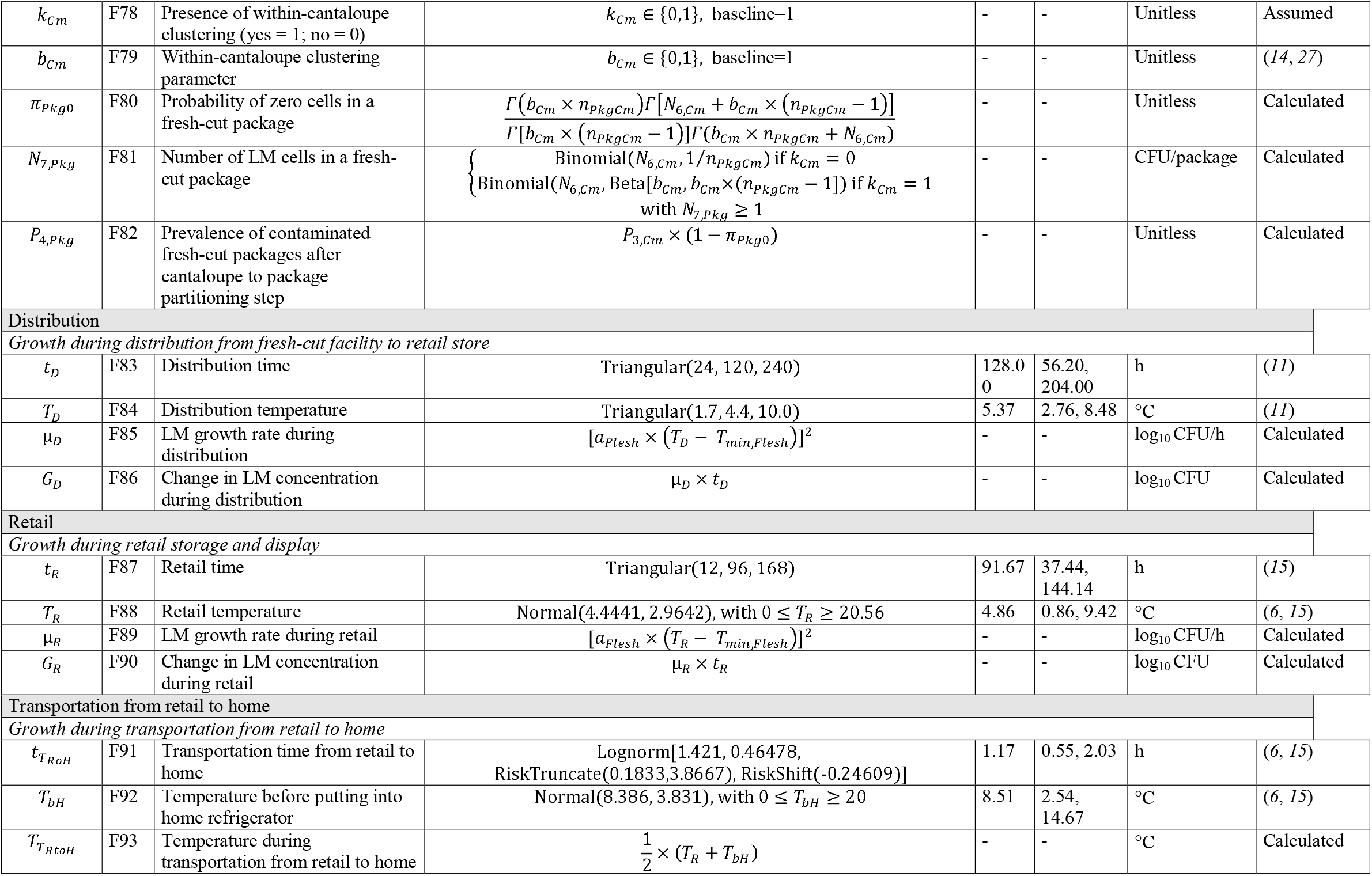
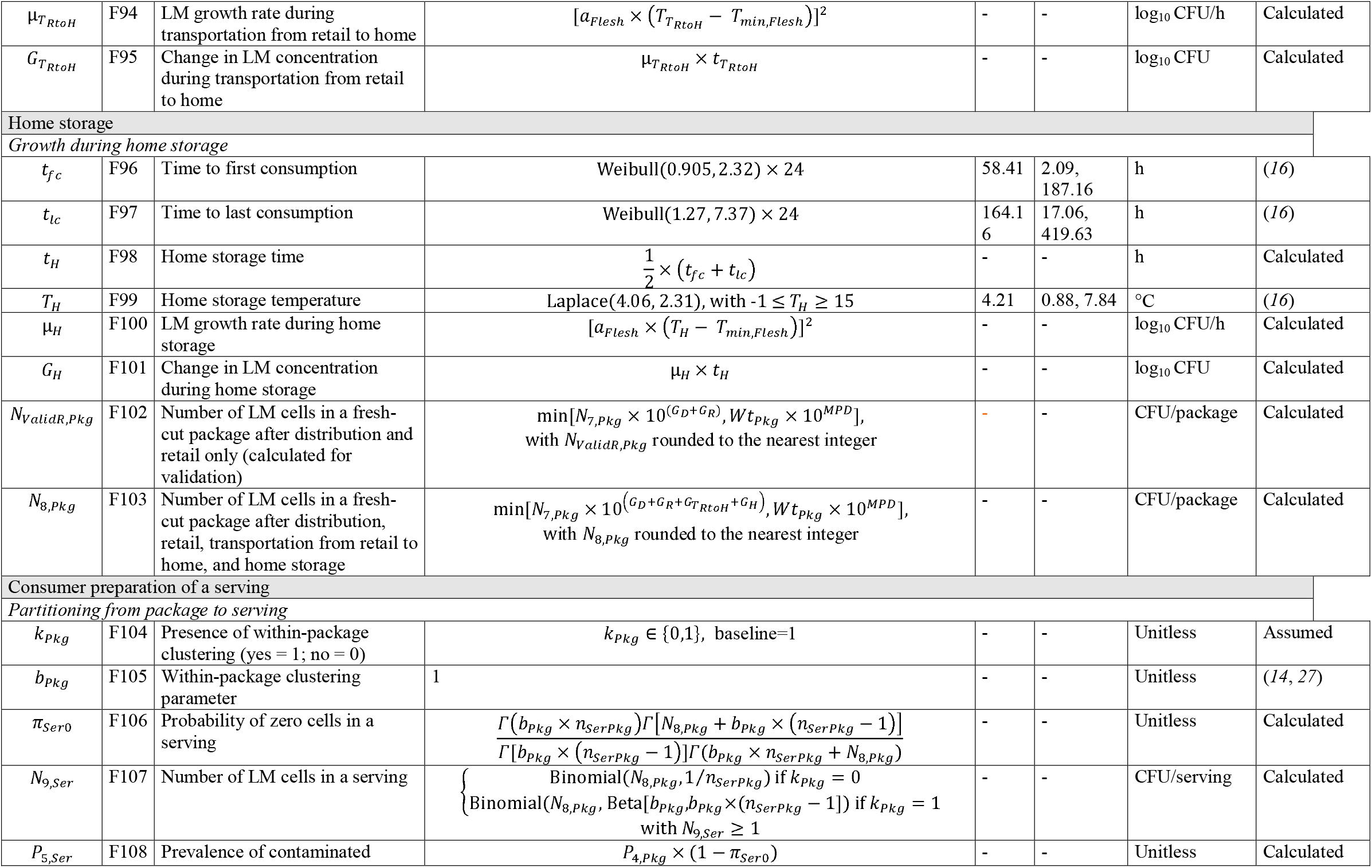
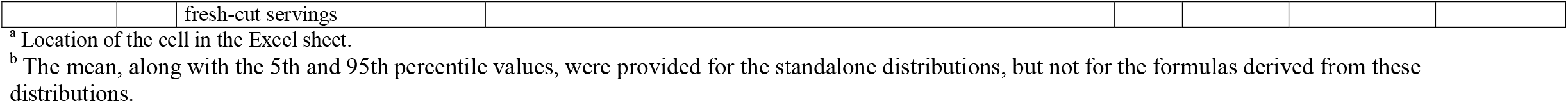
Model parameters and their sources.

**Table 2.**
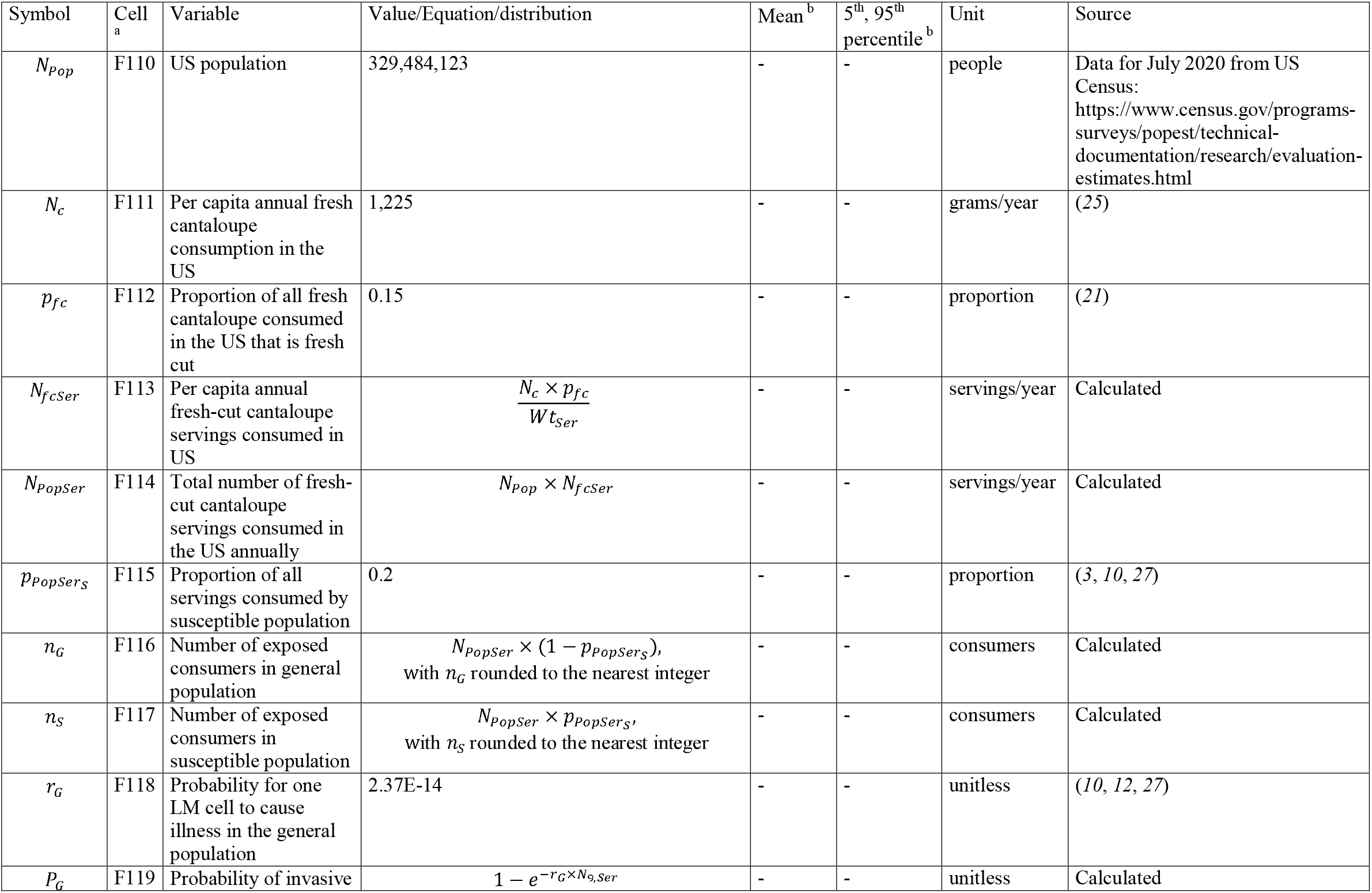
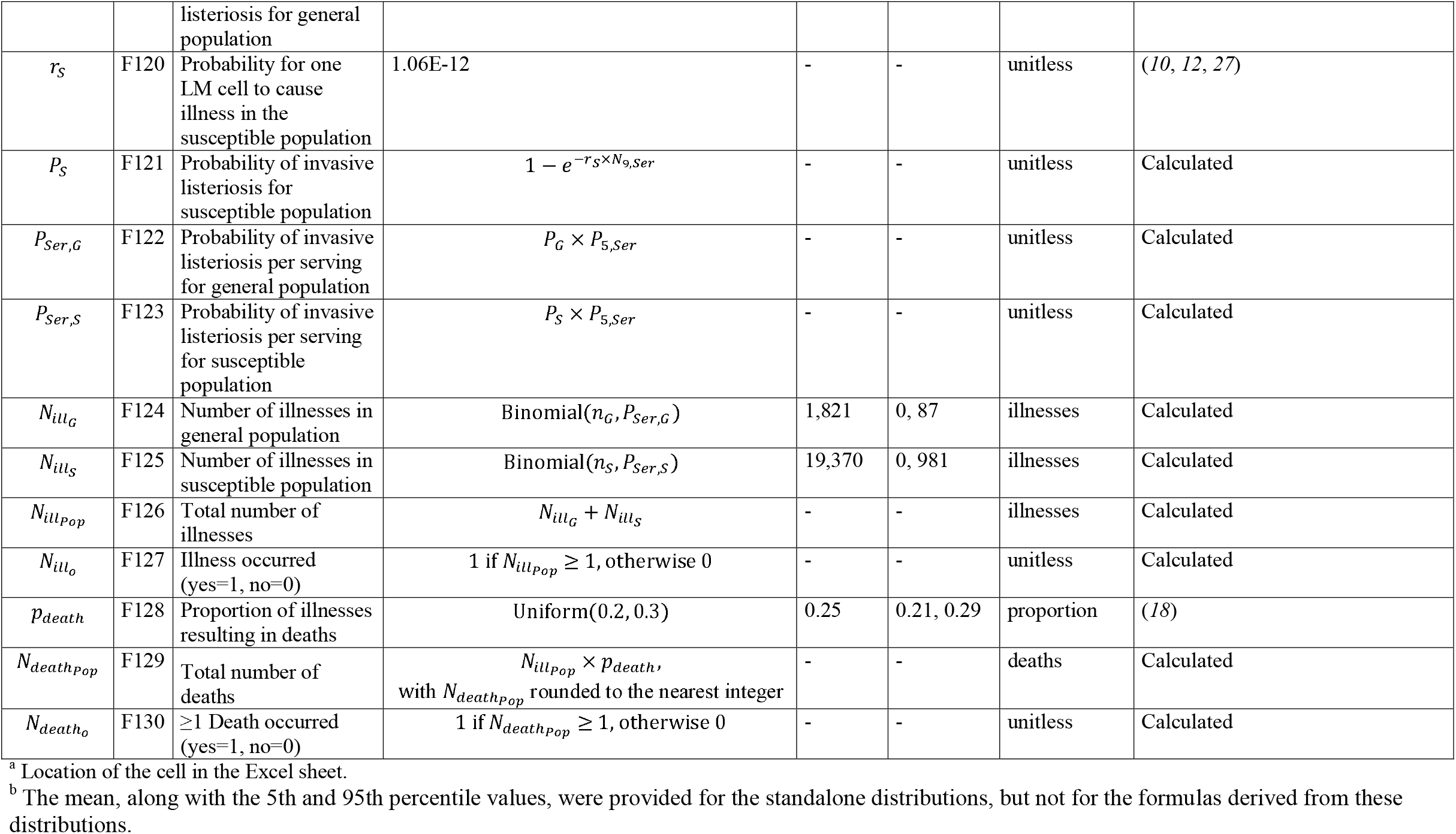
Exposure settings predicting model outputs.

| Symbol | Cell<sup>a</sup> | Variable | Value/Equation/distribution | Mean<sup>b</sup> | 5<sup>th</sup>, 95<sup>th</sup><br>percentile<sup>b</sup> | Unit | Source |
| --- | --- | --- | --- | --- | --- | --- | --- |
| $N_{Pop}$ | F110 | US population | 329,484,123 | - | - | people | Data for July 2020 from US Census:<br>https://www.census.gov/programs-surveys/popest/technical-documentation/research/evaluation-estimates.html |
| $N_c$ | F111 | Per capita annual fresh cantaloupe consumption in the US | 1,225 | - | - | grams/year | (25) |
| $p_{fc}$ | F112 | Proportion of all fresh cantaloupe consumed in the US that is fresh cut | 0.15 | - | - | proportion | (21) |
| $N_{fcSer}$ | F113 | Per capita annual fresh-cut cantaloupe servings consumed in US | $\frac{N_c \times p_{fc}}{Wt_{Ser}}$ | - | - | servings/year | Calculated |
| $N_{PopSer}$ | F114 | Total number of fresh-cut cantaloupe servings consumed in the US annually | $N_{Pop} \times N_{fcSer}$ | - | - | servings/year | Calculated |
| $p_{PopSerS}$ | F115 | Proportion of all servings consumed by susceptible population | 0.2 | - | - | proportion | (3, 10, 27) |
| $n_G$ | F116 | Number of exposed consumers in general population | $N_{PopSer} \times (1 - p_{PopSerS})$,<br>with $n_G$ rounded to the nearest integer | - | - | consumers | Calculated |
| $n_S$ | F117 | Number of exposed consumers in susceptible population | $N_{PopSer} \times p_{PopSerS}$,<br>with $n_S$ rounded to the nearest integer | - | - | consumers | Calculated |
| $r_G$ | F118 | Probability for one LM cell to cause illness in the general population | 2.37E-14 | - | - | unitless | (10, 12, 27) |
| $P_G$ | F119 | Probability of invasive listeriosis for general population | $1 - e^{-r_G \times N_{9,Ser}}$ | - | - | unitless | Calculated |
| $r_s$ | F120 | Probability for one LM cell to cause illness in the susceptible population | 1.06E-12 | - | - | unitless | (10, 12, 27) |
| $P_s$ | F121 | Probability of invasive listeriosis for susceptible population | $1 - e^{-r_s \times N_{9, Ser}}$ | - | - | unitless | Calculated |
| $P_{Ser, G}$ | F122 | Probability of invasive listeriosis per serving for general population | $P_G \times P_{5, Ser}$ | - | - | unitless | Calculated |
| $P_{Ser, S}$ | F123 | Probability of invasive listeriosis per serving for susceptible population | $P_S \times P_{5, Ser}$ | - | - | unitless | Calculated |
| $N_{ill, G}$ | F124 | Number of illnesses in general population | $\text{Binomial}(n_G, P_{Ser, G})$ | 1,821 | 0, 87 | illnesses | Calculated |
| $N_{ill, S}$ | F125 | Number of illnesses in susceptible population | $\text{Binomial}(n_S, P_{Ser, S})$ | 19,370 | 0, 981 | illnesses | Calculated |
| $N_{ill, Pop}$ | F126 | Total number of illnesses | $N_{ill, G} + N_{ill, S}$ | - | - | illnesses | Calculated |
| $N_{ill, 0}$ | F127 | Illness occurred (yes=1, no=0) | 1 if $N_{ill, Pop} \geq 1$, otherwise 0 | - | - | unitless | Calculated |
| $p_{death}$ | F128 | Proportion of illnesses resulting in deaths | $\text{Uniform}(0.2, 0.3)$ | 0.25 | 0.21, 0.29 | proportion | (18) |
| $N_{death, Pop}$ | F129 | Total number of deaths | $N_{ill, Pop} \times p_{death}$,<br>with $N_{death, Pop}$ rounded to the nearest integer | - | - | deaths | Calculated |
| $N_{death, 0}$ | F130 | $\geq 1$ Death occurred (yes=1, no=0) | 1 if $N_{death, Pop} \geq 1$, otherwise 0 | - | - | unitless | Calculated |

<sup>a</sup> Location of the cell in the Excel sheet.
<sup>b</sup> The mean, along with the 5th and 95th percentile values, were provided for the standalone distributions, but not for the formulas derived from these distributions.

**Figure 1.**
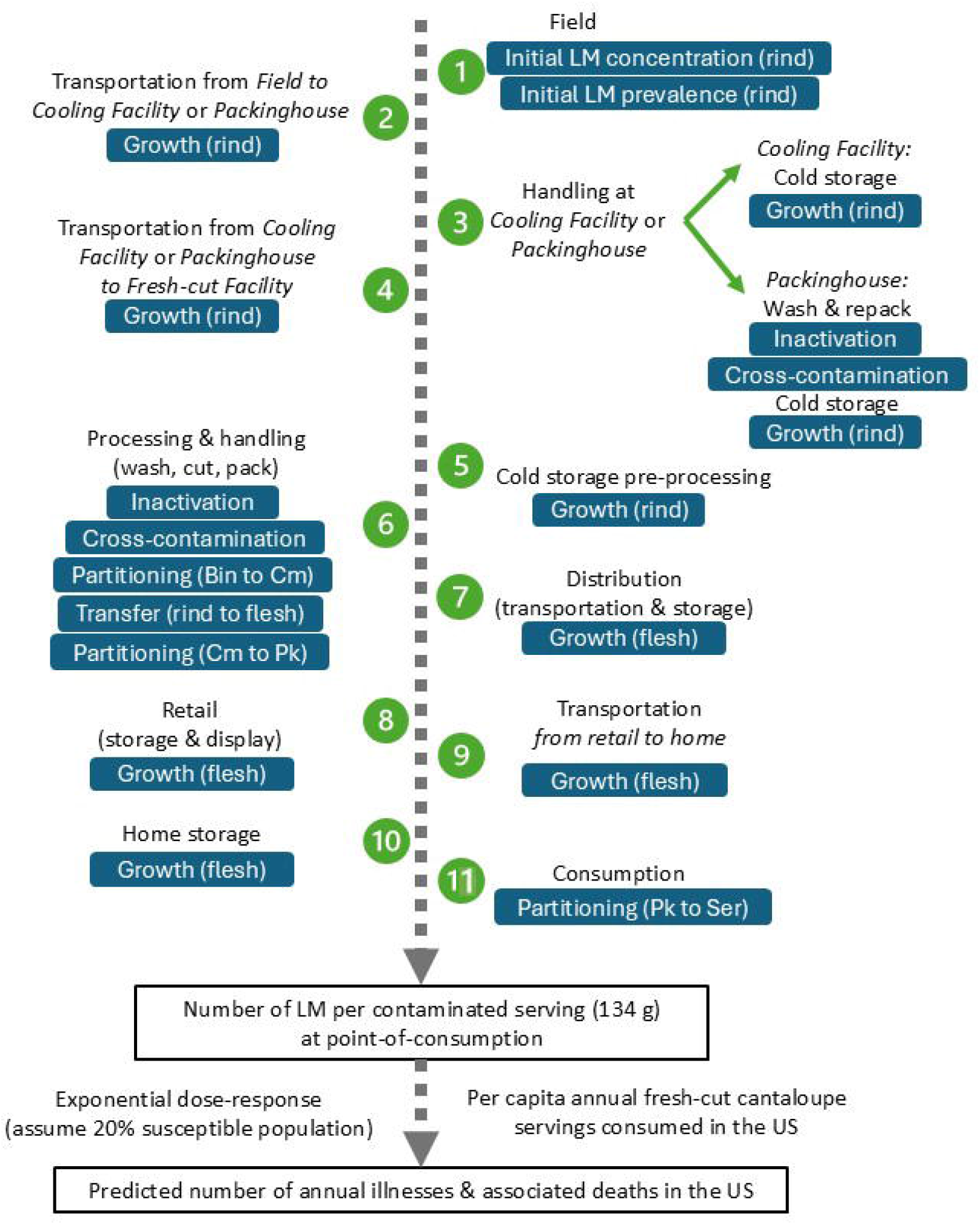
Conceptual model. Green circles indicate module in the model and blue boxes indicate the modeled process. LM: *Listeria monocytogenes*, Cm: individual cantaloupes, Pk: fresh-cut cantaloupe packages and Ser: fresh-cut cantaloupe servings.

### Initial LM contamination and prevalence

The initial probability of a bin of harvested cantaloupes at the field-level containing LM contaminated cantaloupe (*P*_0_,_*Bin*_) was modeled using the Four-parameter Pert(−2.3,-0.6,-0.6,5.4) (implementation on @RISK: BetaSubjective(−2.3,-0.6,(−2.3+-0.6+5.4*-0.6)/(5.4+2),-0.6)) distribution from Sullivan et al. (*33*); this distribution was originally determined from an expert elicitation for prevalence of *Listeria* in unspecified type of produce arriving at a fresh-cut facility and has since been used by Barnett-Neefs et al. (*2*) for the probability of a crate of raw produce containing *Listeria*-contaminated produce upon arrival at a packinghouse.

Due to lack of any data on the levels of LM on cantaloupes at the field-level, the initial number of LM cells on the total cantaloupe surface area in a contaminated bin (*N*_0_,_*Bin*_) was determined as follows

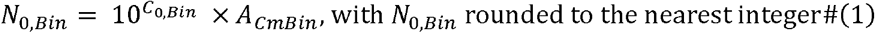

where *C*_0_,_*Bin*_ is the initial concentration of LM on the surface area of cantaloupes in a contaminated bin (log_10_ CFU/cm^2^) and *A*_*CmBin*_ is the total cantaloupe surface area in a bin (cm^2^). Throughout the model, the minimum number of LM per contaminated unit (i.e., bin, cantaloupe, package, or serving) was set at 1 CFU and rounding was applied when necessary to prevent partial LM cells. The assumption of a minimum of 1 CFU per unit was a limitation with negligible impact, affecting 3.8 % of the iterations. The maximum population density (*MPD)* for LM was set at 9 log_10_ CFU/g or CFU/cm^2^, which has been used in previous studies (*13, 15*). Based on this, along with an assumption that the most likely initial LM concentration was -1 log_10_ CFU/cm^2^, *C*_0_,_*Bin*_ was determined as follows

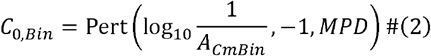

*A*_*CmBin*_ was estimated as follows

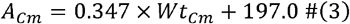

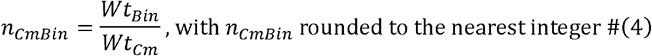

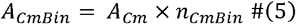

where *A*_*Cm*_ is the mean surface area of a cantaloupe (cm) which was calculated using an equation from Eifert et al. (*11*). *n*_*CmBin*_ is the number of cantaloupes in a bin given the total weight of cantaloupes in a bin, *Wt*_*Bin*_, which was set to 454,000 g (1/2 ton), and the weight of a cantaloupe, *Wt*_*Cm*_, which was set to 1,360 g (3 lbs).

There have not been previous reports of field-level differences in LM contamination for produce destined for either a cooling facility or packinghouse, therefore the initial LM concentration or prevalence for Scheme A (field → cooling facility → fresh-cut facility) or B (field → packinghouse → fresh-cut facility) bins were assumed to be the same. Due to lack of available data on industry-wide practices, it was also assumed for the baseline that 50% of the cantaloupe supply followed Scheme A (*p*_*s*_ = 0.5)

### Cross-contamination in the packinghouse and fresh-cut facility environments

The model included cross-contamination in the packinghouse (*i* = *pk)* and fresh-cut facility (*i* = *Fc)* environments, where LM on cantaloupes could be transferred to food contact surfaces (FCS) and LM on FCS could be transferred to cantaloupes during packing or processing. It was assumed that cross-contamination did not occur at the cooling facility, given cantaloupes remained in the field-packed bin. The effect of cross-contamination on the redistribution of LM among cantaloupes was not modeled because it would not affect the total number of LM cells on cantaloupes in the bin.

The number of LM cells initially present on contaminated 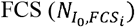 at stage *i*(*i* = *pk* or *Fc)* was modeled using the Four-parameter Pert(0.2,3.3,3.7,3.3) distribution adopted from an agent based model of *Listeria* in a fresh-cut facility (*33*), where the distribution was obtained through expert elicitation to describe the number of *Listeria* cells introduced onto FCSs through unpredictable random events (e.g. maintenance). It was assumed that this distribution is an acceptable representation of the FCS contamination from a previous shift. Cleaning and sanitation of the FCS before the shift was modeled as inactivation, based on the “Method #3” approach (explained below) for inactivation of *D*log_10_ cfu from Pouillot et al. (*27*), which is an alternative to their “Method #2” approach of

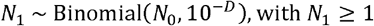

that can be difficult to simulate in certain situations such as when *N*_0_ is small and 10^−*D*^ is close to 0. 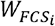 is defined as the status of cleaning and sanitation of FCS (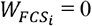, no cleaning and sanitation; 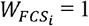, cleaning and sanitation; 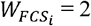, cleaning only), assuming these activities occurred before the shift. The baseline assumed no cleaning and sanitation of FCS occurred 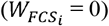. As part of the implementation of the “Method #3” (*26*), given 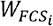, the number of LM cells initially on contaminated FCS equipment after any cleaning and sanitation 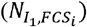 was as follows

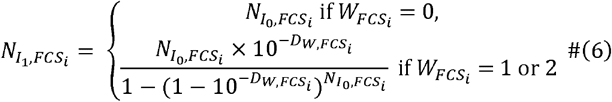

where 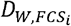 is the expected log reduction of LM due to inactivation by the selected FCS cleaning and sanitation. The 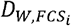 for 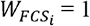 (i.e., cleaning and sanitation) was modeled as a Four-parameter Pert(−8,-6,-1.5,4) distribution and for 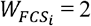 (i.e., cleaning only) was modeled as a Four-parameter Pert(−1.5,-0.5,0,4) distribution from Barnett-Neefs et al. (*2*) and Gallagher et al. (*16*).

Based on the approach from Gallagher et al. (*16*), cross-contamination of LM in the environment at stage *i*(*i* = *Pk* or *Fc)* was modeled as

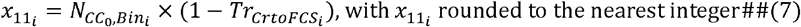

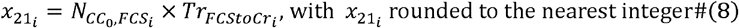

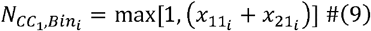

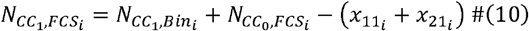

where 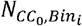 is the initial number of LM cells on cantaloupe rinds in bin and 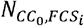 (i.e., 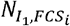 is the initial number of LM cells on FCS, before cross-contamination. 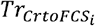 is the transfer coefficient from cantaloupe rinds to 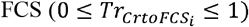 which was modeled using a Normal(−0.28,0.2) distribution from Hoelzer et al. (*18*) and 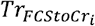 is the transfer coefficient for LM transmission from FCS to cantaloupe rinds 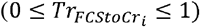, which was modeled using a Normal(−0.44,0.4) distribution from Hoelzer et al. (*18*). 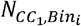 is the number of LM cells on cantaloupe rinds in bin and 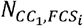 is the number of LM cells on FCS, after cross-contamination. The prevalence of contaminated cantaloupe units after the cross-contamination step 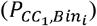 was determined using a simplistic approach from Pang et al. (*26*), as follows

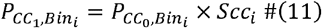

where 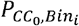 is the prevalence of contaminated bins before cross-contamination and *Scci* is the spread of contamination due to cross-contamination, which was modeled as a Pert(1,1.2,2) distribution from Pang et al. (*26*).

### Washing

The model included washing of whole cantaloupes at the packinghouse (*i* = *Pk)* and fresh-cut facility (*i* = *Fc)*, simulated as inactivation for an individual bin of cantaloupes, assuming the inactivation is applied homogenously and independently on each LM cell on the surfaces of cantaloupes in the bin (*23, 27*). The approach used for modeling inactivation due to wash was based on the aforementioned “Method #3” approach from Pouillot et al. (*27*). As such, the number of LM cells on the surfaces of cantaloupes in the bin after inactivation 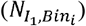 was determined using the model

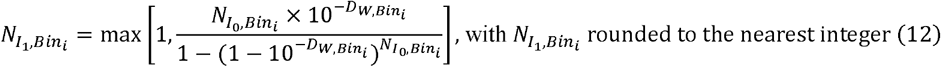

where 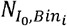 is the number of LM cells on the unit (bin of cantaloupes) before inactivation (CFU/unit) and 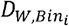 is the expected log reduction of LM due to inactivation by wash. For 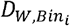 a Pert(0.01,1,3) distribution was used to represent the expected log reduction across currently used post-harvest sanitation methods using industry relevant parameters (e.g., ≤ 2 min contact time) as reported in Bartlett et al. (*3*); sanitation methods modelled included: chlorine, PAA, and chlorine dioxide (aqueous), and water only (i.e., water rinse without any sanitizer). The prevalence of contaminated bins after inactivation 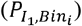 was determined using the model

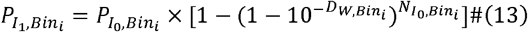

where 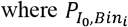 is the prevalence of contaminated bins before inactivation.

### Cutting

Transfer of LM cells from cantaloupe rind to flesh during the cutting step was modeled as

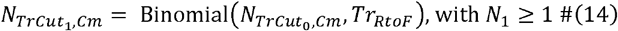

where 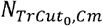 is the number of LM cells on the cantaloupe rind of the individual cantaloupe before cutting, 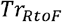 is the transfer coefficient for LM transmission from cantaloupe rind to flesh during cutting, and 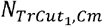 is the number of LM cells on cantaloupe flesh after cutting. The transfer coefficient, 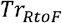 is modeled using a Uniform(0.01,0.0631) distribution using data from two previous studies (*41, 42*).

### Partitioning from bins to individual cantaloupes to packages to servings

Partitioning, splitting of a unit into smaller subunits, occurred 3 times in the model: (i) step = processing at fresh-cut facility, unit = bin, subunit = cantaloupe; (ii) step = packing at fresh-cut facility, unit = cantaloupe, subunit = package; and (iii) step = consumer preparation, unit = package, subunit = serving. The respective models for partitioning of the units into subunits considered within-unit clustering of LM cells due to clumping or aggregation of cells, using the approach from Nauta (*24*) and Zoellner et al. (*46*). For each step *j*, partitioning from *unit*_*j*_ to *subunit*_*j*_ was modeled as follows

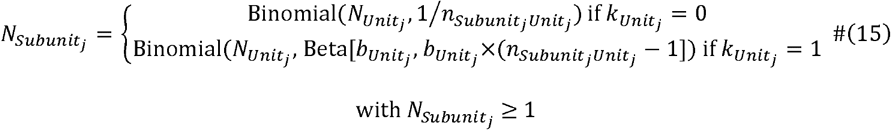

where 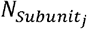 is the number of LM cells in a subunit after partitioning, 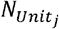 is the number of LM cells in a unit before partitioning, and 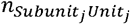 represents the number of subunits originating from a given unit. The parameter 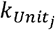 represents presence of clustering within a unit (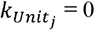, no clustering; 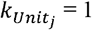, clustering) and 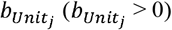 represents within-package clustering, where 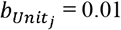 indicates highly clustered contamination and 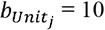 indicates homogenous contamination within a contaminated unit. Following the approach used by Zoellner et al. (*46*), a baseline of 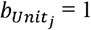 was assumed, representing a moderate level of clustering with different 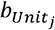 values tested in the scenario analysis. The probability that a subunit originating from a unit containing *N* cells is not contaminated 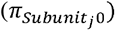 was modeled as follows

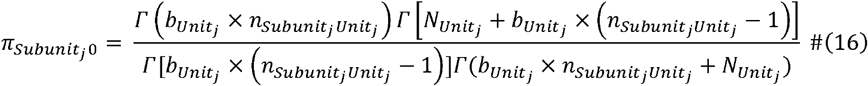

where Γ is the gamma function (*24, 46*). The prevalence of contaminated subunits after partitioning 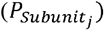 was determined using the model

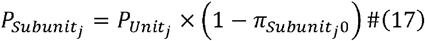

where 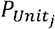 is the prevalence of contaminated units before partitioning.

### Time-temperature conditions across supply chain steps

Transportation temperature (°C) from the field to the intermediate facility 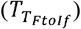 was modeled using a Uniform(21,35) distribution based on the range reported by Fairbank et al. (*12*) for cantaloupe temperatures after transportation from field to a packinghouse in California. A Triangular(0.5,12,48) distribution was used for the transportation time (h) from the field to the intermediate facility 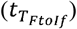 based on input from experts (1 industry professional, 2 academics). Temperature (°C) for storage at the intermediate facility 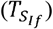, transportation from intermediate facility to fresh-cut facility 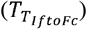, and storage pre-processing at the fresh-cut facility 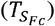, were modeled using a Uniform(2,6) distribution based on the recommended temperature range from the FDA’s National Commodity-Specific Food Safety Guidelines for Cantaloupe and Netted Melons (*40*). Time (h) for storage at the intermediate facility 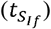 was modeled as a Triangular(1,12,24) distribution, transportation from intermediate facility to fresh-cut facility 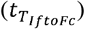 was modeled as a Triangular(0.5,12,48) distribution, and storage pre-processing at fresh-cut facility 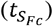 was modeled as a Triangular(1,12,24) distribution based on input from experts (1 industry professional, 2 academics). Distribution time (h) (*t*_*D*_) was modeled using a Triangular(24,120,240) distribution and temperature (°C) (*T*_*D*_) was modeled as a Triangular(1.7,4.4,10.0) distribution, as reported in the 2015 Joint FDA/Health Canada LM soft-ripened cheese risk assessment (*15*). Retail storage time (h) (*t*_*R*_) was modeled using a Triangular(12,96,168) distribution from Pang et al. (*26*) for fresh-cut lettuce. Retail storage temperature (*T*_*R*_) was modeled using a Normal(4.4441,2.9642) distribution truncated at a minimum of 0 and maximum of 20.56°C from Pang et al. (*26*) based on retail refrigerated display data from the EcoSure cold temperature report (*10*). Temperature during transportation from retail to home 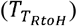 was modeled based on the approach used by Latorre et al. (*19*) and Pang et al. (*26*) from the EcoSure report (*10*) as follows:

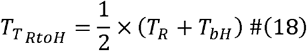

where *T*_*bH*_ is the temperature before putting the package into home refrigerator, modeled using a Normal(8.386,3.831) distribution truncated at a minimum of 0 and maximum of 20°C based on Pang et al. (*26*) and the EcoSure report (*10*). Home storage temperature (*T*_*H*_) was modeled using the Laplace(4.06,2.31) distribution from Pouillot et al. (*28*), which we adapted to include truncation at -5 and 13°C. Transportation time (h) from retail to home 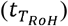 was modeled using the

Lognormal(1.421,0.46478) distribution truncated at 0.1833 and 3.8667 with Shift(−0.24609) from Pang et al. (*26*), which had been fitted using data from the EcoSure report (*10*). Home storage time (h) (*t*_*H*_) was modeled as

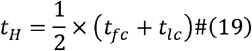

where *t*_*fc*_is the time to first consumption modeled as a Weibull(0.905,2.32) distribution and *t*_*lc*_ is the time to last consumption modeled as a Weibull(1.27,7.37) distribution from Pouillot et al. (*28*) for precut fresh fruit. Temperature was assumed to be constant within each step of the model.

### Microbial growth kinetics of *Listeria monocytogenes* on cantaloupe

The growth model for LM developed by Topalcengiz et al. (*35*) was used to model the change in LM concentration on cantaloupe rinds. To our knowledge, no other studies modeling LM growth on cantaloupe rinds have been published. The growth model for LM developed by Danyluk et al. (*7*) was used to model the change in cantaloupe concentration on cantaloupe flesh in this study. Among the available growth models, the Danyluk et al. (*7*) model was selected based on the methodology (e.g., experimental design included temperatures from 4 to 25°C) and demonstration of model accuracy relative to a similar study by Fang et al. (*13*). Growth of LM on cantaloupe matrix *k* was determined using the primary model

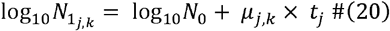

where *N*_*j,k*_ is the concentration at step *j* (*k* rind, CFU/cm^2^ ; *k* = flesh, CFU/g), μ_*j,k*_ is the growth rate at a specific temperature (*k* = rind, log_10_ CFU/cm^2^/h; *k* = flesh, log_10_ CFU/g/h), *t*_*j*_ is time at step *j*, and *N*_0_ is the concentration at time 0 (CFU/cm^2^ or CFU/g). Growth rate at a specific temperature (*μ*_*j,k*_) was determined using the secondary model

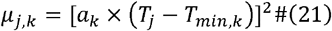

where *a*_*k*_ is the temperature coefficient (*k* = rind, 0.005; *k* = flesh, 0.0186), _j_ is the temperature (°C) at step *j*, and *T*_*min,k*_ is the theoretical minimum growth temperature in °C (*k* = rind, -1.4; *k* = flesh, -0.5108) (*7, 35*).

### Dose-response modeling and risk characterization

The risk of listeriosis per serving was predicted from the estimated dose of contamination consumed in a single serving of fresh-cut cantaloupe. Two subpopulations of consumers, the susceptible population and the general population, were considered. The exponential dose-response model and parameters used in previous risk assessment models of LM contamination in foods (*14, 16, 46*) were applied to estimate probability of listeriosis from consumption of a LM-contaminated fresh-cut cantaloupe serving, as follows:

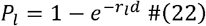

where *P*_*l*_ is the probability of listeriosis in subpopulation *l* given an individual exposure to LM from a dose *d* (CFU) and *r*_*l*_ the probability that one LM cell will cause illness (*l =* general, 2.37E-14; *l =* susceptible, 1.06E-12). The probability of invasive listeriosis per serving in subpopulation *l* (*P*_*ser,l*_)was calculated using *P*_*l*_ and the prevalence of contaminated servings (*P*_*ser*_):

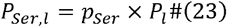

The predicted total number of illnesses and deaths attributed to LM-contaminated fresh-cut cantaloupe in the US annually were determined. First, the total number of fresh-cut cantaloupe servings consumed in the US annually (*N*_*PopSer*_) was estimated as follows:

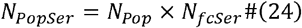

where *N*_*pop*_ is the total number of people in the US population, which was set to 329,484,123 people based on US Census data from July 2020 (*38*) and *N*_*fcser*_ is the per capita fresh-cut cantaloupe consumption in the US (servings/year) which was estimated as follows:

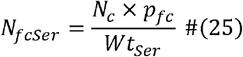

where *N*_*c*_ is the per capita fresh cantaloupe consumption in the US (g/year), which was set to 1225 g/year based on the estimated 2018 per capita edible weight for fresh cantaloupe from the US Department of Agriculture Economic Research Service (*43*), *p*_*fc*_ is the proportion of fresh cantaloupe consumed in the US that is fresh-cut, which was set to 15% based on Torres et al. (*36*), and *Wt*_*Ser*_ is the weight of a single fresh-cut cantaloupe serving, which was set to 134 g based on (*39*). Multiple servings consumed by the same consumer were considered independent of each other. Consistent with previous studies, the proportion of all servings consumed by the susceptible population 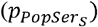 was assumed to be 0.2 (*4, 14, 46*). Thus, the number of exposed consumers in subpopulation *l*(*n*_*l*_) was:

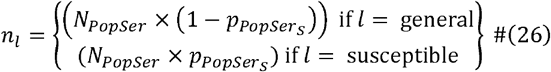

The number of illnesses in subpopulation *l* was modeled as:

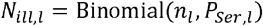

The total number of illnesses, 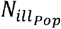, was calculated as follows:

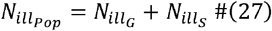

Given the proportion of illnesses resulting in deaths (*p*_*death*_), modeled as Uniform(0.20,0.30) based on the reported mortality rate of 20 to 30% from Swaminathan and Gerner-Smidt (*34*), the total number of deaths, 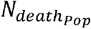, was calculated as follows:

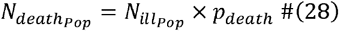

The probability of ≥1 illness and ≥1 death annually attributed to fresh-cut cantaloupe were calculated by counting iterations where illnesses or deaths were ≥1 across all iterations.

### Model validation

For model validation, estimates were compared to the reported illnesses and deaths from the 2011 US listeriosis outbreak (*21*), which is the only recorded LM outbreak attributed to melons in the US (*5, 44*). Additionally, model estimates for LM levels at retail were compared to available data from a market basket survey conducted in the US from January to October 2014 (*45*).

### Scenario analysis

Potential interventions across the supply chain were determined based on literature review and facilitated discussions, led by author SIM, with the study co-authors and with industry experts serving on an advisory council for USDA-NIFA-SCRI grant #2019-51181-30016. To gather further information about industry practices and potential interventions, three informal interviews were also conducted with experts from produce production and processing, distribution and retail, and sanitation industries, respectively. Additionally, a survey was prepared in Qualtrics (Provo, UT) and administered to the project advisory council to identify and rank intervention strategies; in total, there were 12 respondents. Based on the survey results, literature review, and the capability of the developed model, a set of interventions were selected for the scenario analysis (Table 3, Table 4). Clustering parameters (*b*_*Bin*_, *b*_*Cm*_, *b*_*Pkg*_) and *p*_*s*_ were also assessed as part of the scenario analysis (Table 3).

**Table 3.** Parameters and values considered in the scenario analysis.

| Symbol(s) | Parameter | Baseline value | Alternative values |
| --- | --- | --- | --- |
| $p_s$ | Proportion following Scheme A (Field → Cooling facility → Fresh-cut facility) rather than Scheme B (Field → Packinghouse → Fresh-cut facility) | 0.5 | 0; 0.25; 0.75; 1 |
| $W_{FCS,Pk}$ | Status of FCS cleaning and sanitation schedule, LM log reduction on FCS due to scheduled cleaning and sanitation at Packinghouse | No recent FCS cleaning and sanitation:<br>$W_{FCS,Pk} = 0$ | Recent FCS cleaning & sanitation:<br>$W_{FCS,Pk} = 1, D_{W_{FCS,Pk}} = \text{Pert}(-8, -6, -1.5, 4)$;<br>Recent FCS cleaning only:<br>$W_{FCS,Pk} = 2, D_{W_{FCS,Pk}} = \text{Pert}(-1.5, -0.5, 0, 4)$ |
| $W_{FCS,Fc}$ | Status of FCS cleaning and sanitation schedule, LM log reduction on FCS due to scheduled cleaning and sanitation at Fresh-cut facility | No recent FCS cleaning and sanitation:<br>$W_{FCS,Fc} = 0$ | Recent FCS cleaning & sanitation:<br>$W_{FCS,Fc} = 1, D_{W_{FCS,Fc}} = \text{Pert}(-8, -6, -1.5, 4)$;<br>Recent FCS cleaning only:<br>$W_{FCS,Fc} = 2, D_{W_{FCS,Fc}} = \text{Pert}(-1.5, -0.5, 0, 4)$ |
| $t_{SIf}$ | Storage time at Cooling facility or Packinghouse (hours) | Triangular(1,12,24) | Extended storage time (e.g., due to supply chain disruption):<br>Triangular(1,12,48) |
| $t_D, t_R, t_H$<br>$T_D, T_R, T_H$ | Time (hours) and temperature (°C) at Distribution, Retail storage/display, and Home storage | See Table 1 | Reduce $T_D, T_R, T_H$ each by 10%;<br>Reduce $t_D, t_R, t_H$ each by 10%;<br>Reduce $T_D, T_R, T_H, t_D, t_R, t_H$ each by 10% |
| $b_{Bin}$ | Within-bin clustering parameter (partitioning: bin → individual cantaloupe) | 1 | 0.01; 0.1; 5; 10 |
| $b_{cm}$ | Within-cantaloupe clustering parameter (partitioning: individual cantaloupe → package) | 1 | 0.01; 0.1; 5; 10 |
| $b_{pkg}$ | Within-package clustering parameter (partitioning: package → serving) | 1 | 0.01; 0.1; 5; 10 |

**Table 4.** Parameters and values considered in the additional scenario analysis regarding temperature at Distribution, Retail storage/display, and Home storage.

| Symbol(s) | Parameter | Baseline value | Alternative values |
| --- | --- | --- | --- |
| $T_D$ | Temperature (°C) at Distribution | See Table 1 | Reduce $T_D$ by 10% |
| $T_R$ | Temperature (°C) at Retail storage/display | See Table 1 | Reduce $T_R$ by 10% |
| $T_H$ | Temperature (°C) at Home storage | See Table 1 | Reduce $T_H$ by 10% |
| $T_D$ | Temperature (°C) at Distribution | See Table 1 | Truncate the upper end of $T_D$ from 10.00°C to 7.41°C<sup>a</sup> |
| $T_R$ | Temperature (°C) at Retail storage/display | See Table 1 | Truncate the upper end of $T_R$ from 20.56°C to 7.41°C<sup>a</sup> |
| $T_H$ | Temperature (°C) at Home storage | See Table 1 | Truncate the upper $T_H$ from 15.00°C to 7.41°C<sup>a</sup> |
| $T_D$ | Temperature (°C) at Distribution | See Table 1 | Shift the mean $T_D$ from 4.40°C to 3.00°C<sup>b</sup> |
| $T_R$ | Temperature (°C) at Retail storage/display | See Table 1 | Shift the mean $T_R$ from 4.44°C to 3.00°C<sup>b</sup> |
| $T_H$ | Temperature (°C) at Home storage | See Table 1 | Shift the mean $T_H$ from 4.06°C to 3.00°C<sup>b</sup> |
| $T_D$ | Temperature (°C) at Distribution | See Table 1 | Truncate the upper end of $T_D$ from 10.00°C to 7.41°C<sup>a</sup> and shift the mean $T_D$ from 4.40°C to 3.00°C<sup>b</sup> |
| $T_R$ | Temperature (°C) at Retail storage/display | See Table 1 | Truncate the upper end of $T_R$ from 20.56°C to 7.41°C<sup>a</sup> and shift the mean $T_R$ from 4.44°C to 3.00°C<sup>b</sup> |
| $T_H$ | Temperature (°C) at Home storage | See Table 1 | Truncate the upper $T_H$ from 15.00°C to 7.41°C<sup>a</sup> and shift the mean $T_H$ from 4.06°C to 3.00°C<sup>b</sup> |

<sup>a</sup> 7.41°C (one standard deviation away from the mean retail storage temperature) represents improved refrigeration, chosen by removing the skewed tail of the retail storage temperature distribution. Values beyond one standard deviation were excluded from the distribution to remove the tail. This adjustment, applied for storage during distribution, retail and home storage, reflects a more stringent temperature control of 7.41°C as the maximum, compared to the previous higher value of 20.56°C.
<sup>b</sup> 3.00°C represents refrigerators with a lower average storage temperature.

### Model simulations and analysis

The QMRA model was implemented using @RISK software version 8.6.0 (Lumivero, Denver, CO). For the baseline model and each scenario, the model was simulated for 100,000 iterations with Latin hypercube sampling.

Data analysis and visualization were performed in R Statistical Programming Environment version 4.0.5 (*29*). Partial rank correlation coefficients (PRCC) were used for sensitivity analysis of the baseline model to identify the effect of inputs on key outcomes, performed using the “epiR” R package (*32*) with a significance threshold adjusted using the Bonferroni correction (p = 0.05/28 = 0.0018).

The QMRA model as well as data and code from this study are available on GitHub: [https://github.com/IvanekLab/Cantaloupe_LM_QMRA].

## RESULTS

The baseline model estimated a median of 3.18 log_10_ CFU of LM (5^th^, 95^th^, 99^th^ percentiles: 0.60, 8.06, 11.04) per contaminated serving (134 g) of fresh-cut cantaloupe at consumption (Figure 2A). Under the Exponential does-response model, baseline model estimates for the median (5^th^, 95^th^ percentiles) probability of illness per serving was 1.4 × 10^−12^ (1.8 × 10^−15^, 2.4 × 10^−7^) for general populations and 6.4 × 10^−11^ (7.9 × 10^−14^, 1.1 × 10^−5^) for susceptible populations. The median predicted number of illnesses was 0 (0, 1,070) and deaths was 0 (0, 264). The probability of ≥1 illness and ≥1 death annually attributed to fresh-cut cantaloupe were 24.1% and 18.8%, respectively.

**Figure 2.**
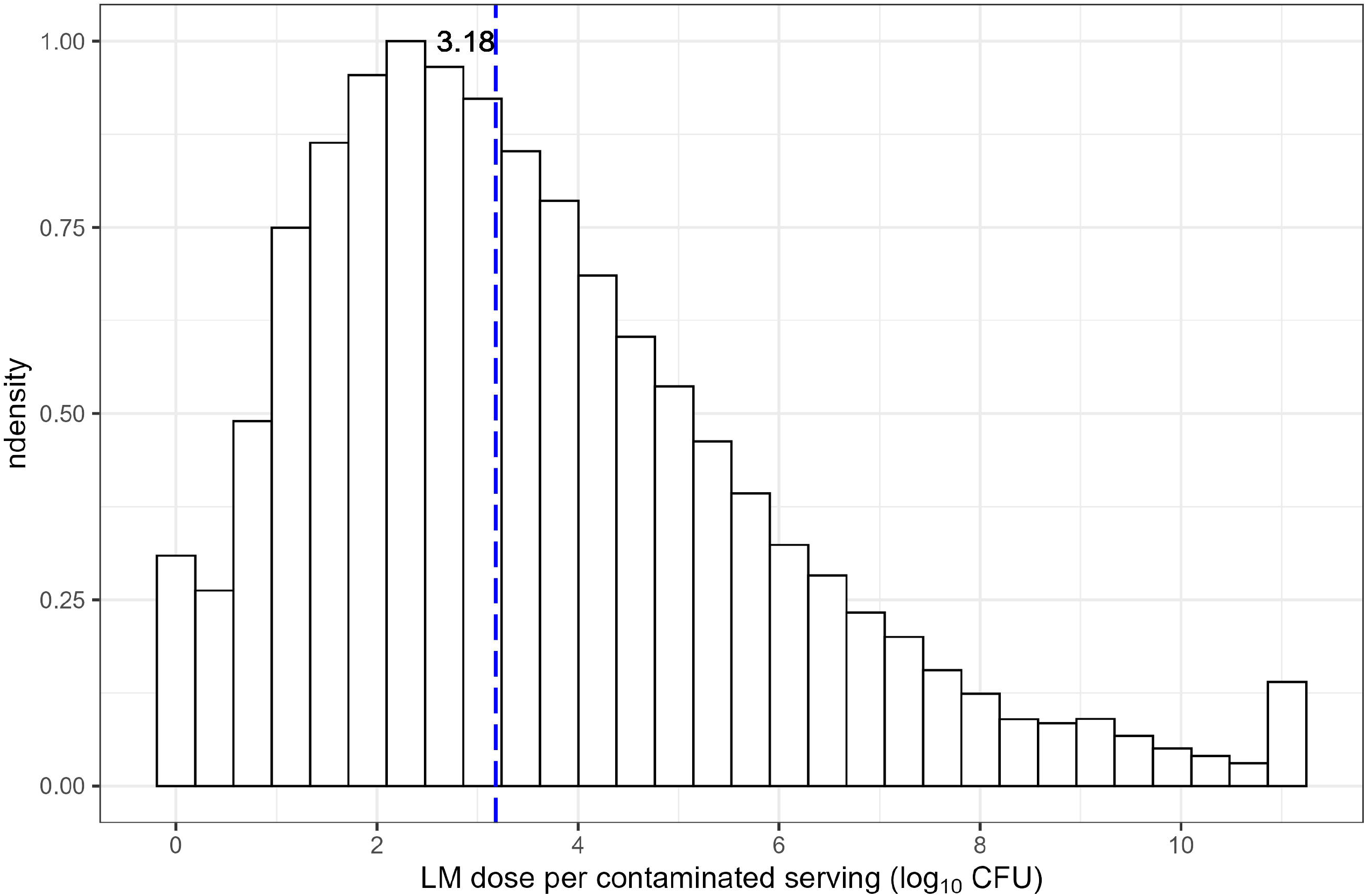
(A) Estimated LM dose (log_10_CFU) per contaminated serving (134 g) of fresh-cut cantaloupe at time of consumption. A maximum of 9 log_10_ CFU/g of LM were set for the model. The vertical blue line indicates the median LM dose per contaminated serving. (B) Ascending cumulative frequency of the predicted number of illnesses due to LM contaminated fresh-cut cantaloupe serving (log_10_); value of 1 was added to allow log transformation when there were 0 illnesses. The vertical red line indicates the recorded number of illnesses from the 2011 US listeriosis outbreak.

Sensitivity analysis identified (i) time and temperature after packaging (i.e., retail, distribution, home storage: *T*_*D*_,*T*_*R*_,*T*_*H*_,*t*_*D*_,*t*_*R*_,*t*_*H*_) and (ii) initial number of LM cells on the total cantaloupe surface area in a bin (*N*_O,*Bin*_) had the greatest impacts on both LM per contaminated fresh-cut cantaloupe serving (log_10_CFU) (Figure 3A) and the number of illnesses annually in the US due to LM-contaminated cantaloupe (Figure 3C). On the other hand, (i) *N*_O,*Bin*_, (ii) initial number of LM cells initially on contaminated FCS at the fresh-cut facility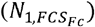, and (iii) transfer coefficients for LM transmission between FCS and cantaloupe at the fresh-cut facility (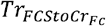 and 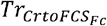) had the greatest impacts on the prevalence of contaminated servings (Figure 3B).

**Figure 3.**
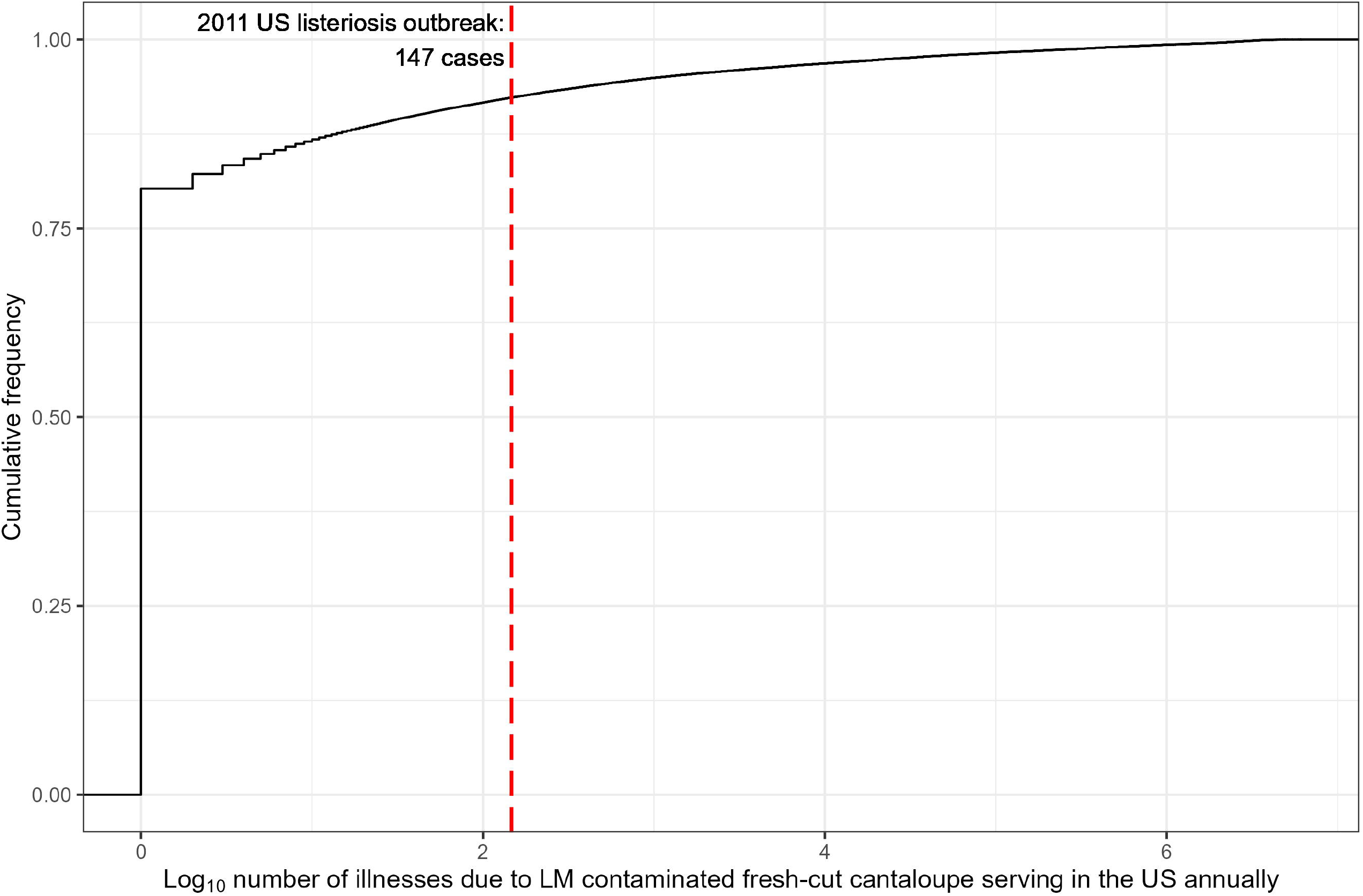
Sensitivity analysis of (A) LM per contaminated fresh-cut cantaloupe serving, (B) prevalence of contaminated servings, and (C) number of illnesses due to LM-contaminated cantaloupe annually in the United States. Bars indicate significant input parameters (P < 0.05/28 after Bonferroni’s correction). Parameters are defined in Table 1.

Scenario analysis showed that the relative degree of clustering affects the risk of listeriosis per serving and the predicted number of illnesses and deaths, where increased clustering (i.e., heterogeneity) of LM contamination generally led to better public health outcomes than homogenous contamination (Figure 4). Assessment of interventions demonstrated that reducing temperature and/or time conditions post-processing can be an effective strategy for reducing LM risk. Increasing the proportion following Scheme A (field → cooling facility → fresh-cut facility) increased LM risk, whereas increasing the proportion following Scheme B (field → packinghouse → fresh-cut facility) decreased LM risk, relative to the baseline of *ps* = 0.5. Specifically, when cantaloupes were processed only through Scheme A, the median (5^th^, 95^th^ percentiles) probability of illness per serving was 2.3 × 10^−12^ (2.1 × 10^−15^, 7.5 × 10^−7^) for general populations and 1.0 × 10^−10^ (9.2 × 10^−14^, 3.3 × 10^−5^) for susceptible populations. When cantaloupes were processed only through Scheme B, the median (5^th^, 95^th^ percentiles) probability of illness per serving was 9.5 × 10^−13^ (1.6 × 10^−15^, 6.9 × 10^−8^) for general populations and 4.2 × 10^−11^ (7.2 × 10^−14^, 3.1 × 10^−6^) for susceptible populations. Among all scenarios evaluated, the log risk of invasive listeriosis per serving from consumption of LM-contaminated cantaloupe was lowest for ‘within package clustering’ parameter, *b*_*pkg*_ = 0.01 and highest for ‘proportion of following Scheme A rather than Scheme B’ parameter, *ps* = 1. Detailed results from the scenario analysis are provided in the Supplemental Material (Supplemental Tables 1 to 8).

**Figure 4.**
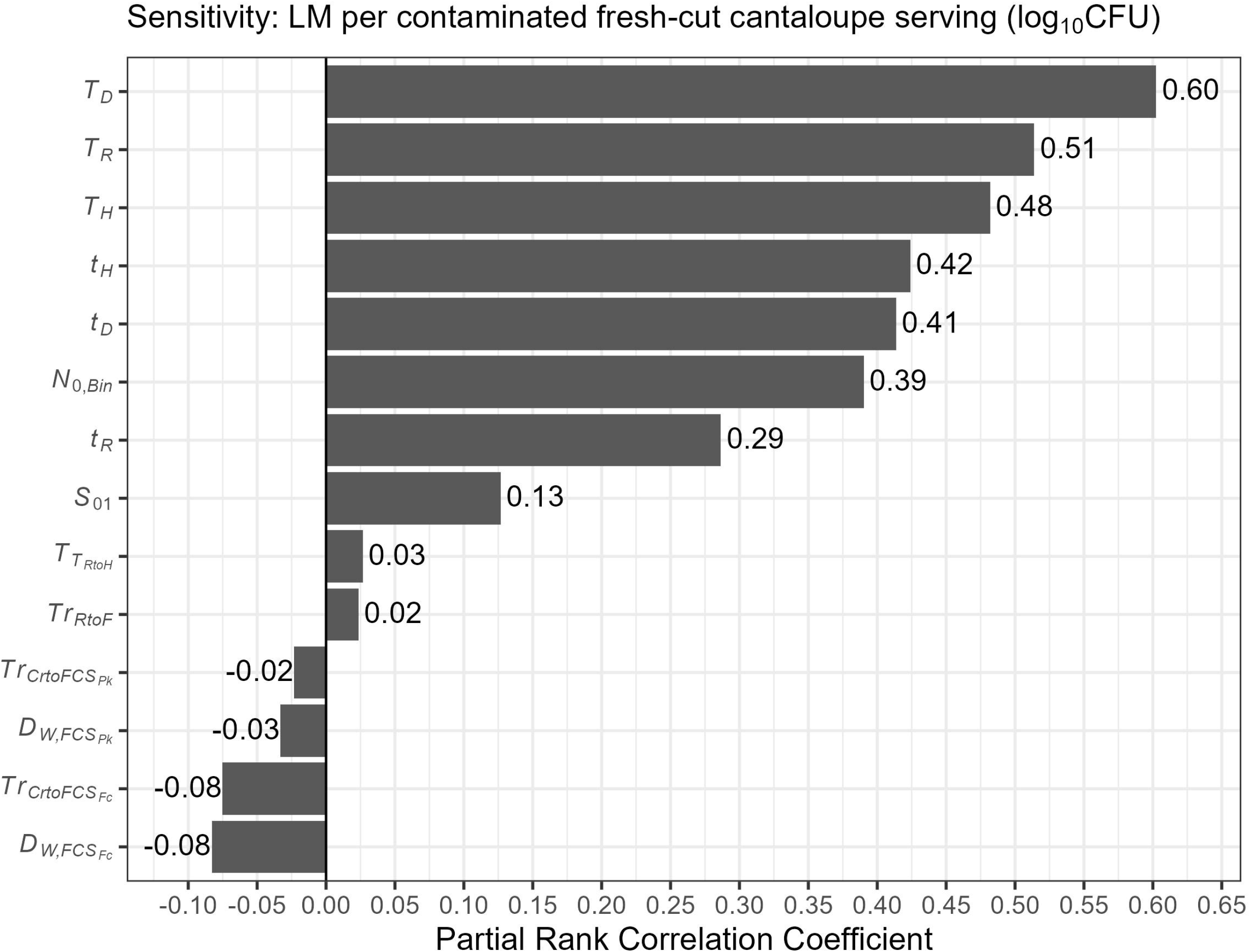
Log risk of invasive listeriosis per serving from consumption of cantaloupe contaminated with *Listeria monocytogenes* according to the exponential dose-response model for general (*P*_*Ser,G*_) and susceptible (*P*_*Ser,S*_) populations, under various scenarios. Violin plots show a combination of a boxplot (median, interquartile range) with a kernel density plot of 100,000 iterations. Red indicates baseline condition. Continuous parameter values were sorted from highest to lowest, while categorical values 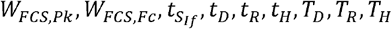 were arranged based on their increasing impact compared to the baseline, with the baseline at the top and the most effective parameter value at the bottom. Parameters and tested scenarios are defined in Table 3.

For model validation, comparison of the predicted levels of LM on fresh-cut cantaloupe at retail from the baseline model to observed levels from a previous study (*6, 45*) showed the observed levels were within the wider range of predicted levels; the observed levels were fit to a normal distribution with a median (5^th^, 95^th^) of -0.97 (−1.53, -0.41), whereas the predicted levels were right skewed with a median of 0.27 (−1.69, 4.46) (Figure 5). Providing further validation of the QMRA model, the recorded number of illnesses and deaths from the 2011 LM-cantaloupe outbreak in the US were each in the 92^nd^ percentile of the estimated annual number of illnesses (Figure 2B) and deaths due to LM contaminated fresh-cut cantaloupe in the US from the baseline model.

**Figure 5.**
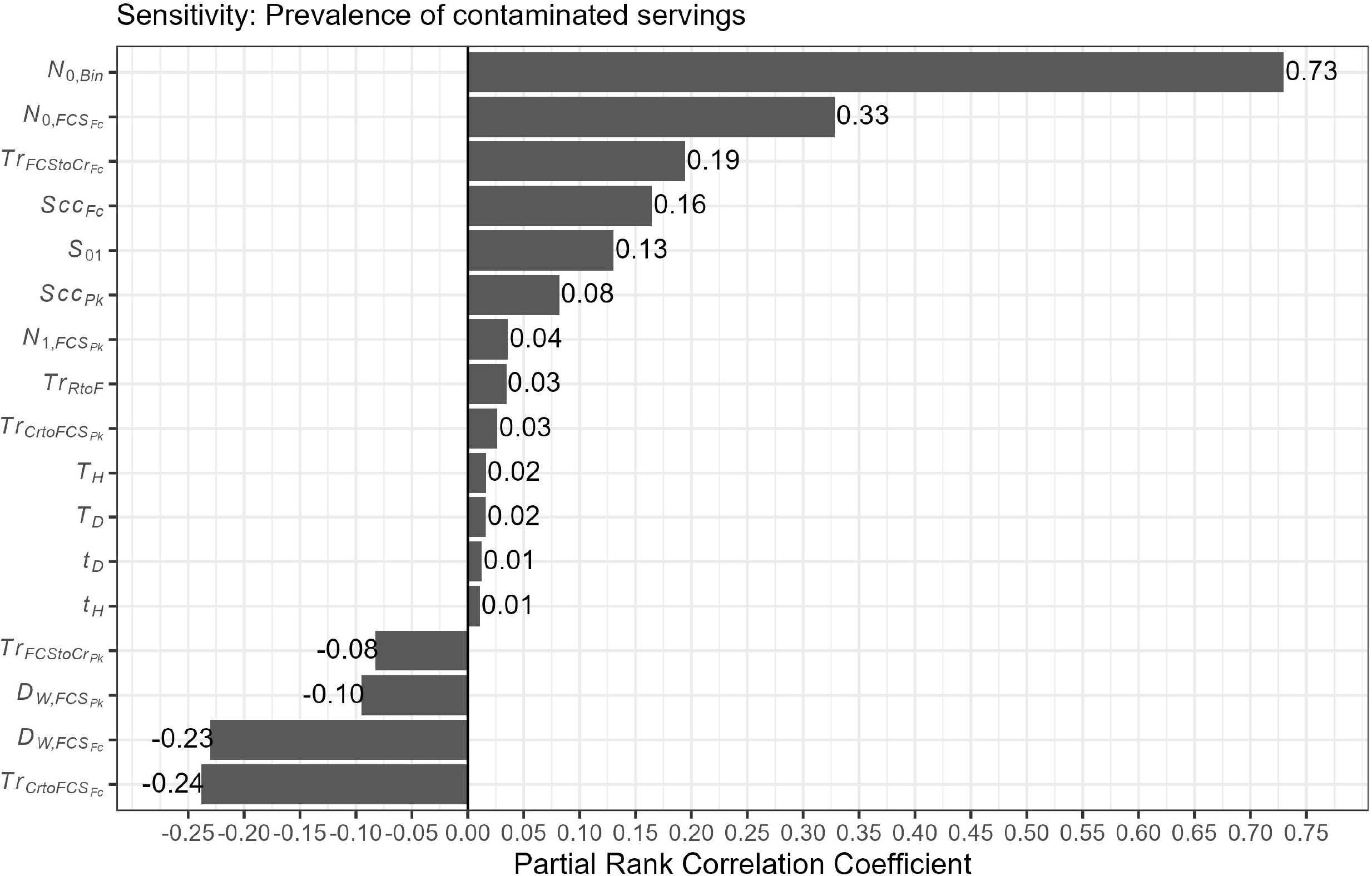
Comparison of predicted levels (indicated in blue) from the baseline model and observed levels (indicated in red) from a previous US retail survey study (*4, 25*) of LM on fresh-cut cantaloupe at retail. The solid line indicates the median and dashed lines indicate 5^th^ (left) and 95^th^ (right) percentiles.

## DISCUSSION

Overall, our findings support that fresh-cut cantaloupe presents a risk to US consumers due to contamination with LM. The QMRA model estimates of annual illnesses and deaths in the US agreed with understanding of occurrence of poorly reported sporadic cases and rare reported large outbreaks (*20*). Our findings support that fresh-cut cantaloupe listeriosis outbreaks are expected to be rare but have the potential to have a major impact under worst-case scenarios. The QMRA model suggests reduction of LM risk to consumers requires development of a multipronged approach aimed at preventing or reducing LM contamination at multiple steps of the fresh-cut supply chain. Importantly, the QMRA model from our study is not intended for immediate use by industry (i.e., cannot be tailored for a specific operation); an operation-specific QMRA would require industry-data for inputs and other parameters. Other limitations, such as lack of knowledge of clustering related parameters, should also be addressed in the future.

Our findings indicated that reducing post-packaging transportation and storage temperatures (distribution, retail, and home storage) may be an effective and practical LM risk reduction strategy. This makes sense as time-temperatures affect the growth and survival of LM (*42*). This finding is consistent with Pang et al. (*26*); a sensitivity analysis of their QMRA model for *E*.*coli* O157:H7 in fresh-cut lettuce identified mean illnesses per year was most sensitive to home storage time, home storage temperature, and *E. coli* concentration in the soil. Findings from the scenario analysis support that reducing time-temperature conditions post-packaging may be an effective strategy for reducing risk of listeriosis due to LM on cantaloupe. Interventions may be designed to improve these conditions and should be assessed for sustainability (economic, social, environmental) in future studies.

Findings from scenario analysis also identified clustering as an important risk factor, where increased clustering generally reduced risk. This finding is consistent with Zoellner et al. (*46*) who also identified that increased clustering reduced the risk of LM on frozen vegetables QMRA. While clustering seems to be important, our understanding is limited. However, an insight for the industry from the clustering scenario is that washing (prior to cutting) that reduces dispersal of LM and cross-contamination also lowers risk of listeriosis. This highlights the importance of washing practices (when used) as part of risk mitigation strategies for cantaloupe food safety.

Due to lack of available data, in order to develop the QMRA model we made a number of assumptions as described in the Methods. The challenges encountered in our study due to lack of available data are consistent with De Keuckelaere et al. (*8*) who summarized data gaps and challenges related to QMRA in fresh produce, including data needs related to (i) prevalence and concentration of pathogens in water or fresh produce along the supply chain, (ii) pathogen transfer during irrigation and washing of fresh and fresh-cut produce, and (iii) reduction, growth, and survival of pathogens along the produce supply chain, and (iv) consumer practices and consumption patterns. Data trusts or other data sharing initiatives should be explored to leverage data already being collected by industry.

Data for LM levels and prevalence in the cantaloupe supply chain were extremely limited, presenting a specific challenge for developing the QMRA described here. While previous studies have investigated LM or *Listeria* spp. on cantaloupe at the field, packinghouse, and processing facility, and retail in the US, positive samples have only been found at the retail level (*37*). This is likely due to limitations of the design of past studies as it is well known that there are sources of LM at the field, packinghouse, and processing facility as demonstrated by previous studies that have detected LM on environmental samples (e.g., soil, water, non-FCS) (*37*). As such, for model validation we used data from a study conducted in the US at the retail level, which detected positive samples and determined LM concentration on the samples (*45*). While retail data are valuable for supply chain modeling, it would be useful, especially for model validation, to have data for comparison at the field-level and at point-of-consumption (i.e., at the beginning & end points of the supply chain). Lack of available data of foodborne pathogen prevalence and concentration is a hurdle for food safety risk assessments and can limit the scope of the model to starting only at the point in the supply chain with available data or require major assumptions.

Our findings indicated the initial concentration of LM on whole cantaloupes at point-of-harvest had a great impact on both LM per contaminated serving and prevalence of contaminated servings. We are unable to assess potential interventions to reduce the initial LM because were unable to include pre-harvest or harvest activities in the QMRA, due to lack of data. For example, our model includes an assumption that there was no difference in LM contamination at the field-level for cantaloupes based on the destined intermediate facility (i.e., packinghouse or cooling facility). While LM prevalence is known to vary across regions in the US (*21*), we accounted for this variability by incorporating a distribution for initial LM counts. However, pathway- and region-specific initial LM counts were not accessed due to limited data availability but should be further explored given the identified impact of the initial LM contamination levels. To allow expansion of the LM-cantaloupe QMRA model to include pre-harvest and harvest activities, studies should be performed at the field level to better understand the sources of and factors affecting LM on cantaloupe at the field (e.g., irrigation), including detection and quantification of LM on environmental sources (e.g., soil or water) to facilitate transmission modeling at the field level.

Another limitation of our study was a simplified representation of cross-contamination. To mechanistically model cross-contamination between facility surfaces and produce would require a model that explicitly considers (i) individual relevant surfaces in a facility (such as in an agent-based model (*33*), and (ii) individual units of produce, which in turn would require data for parameterization and validation of cross-contamination. In the absence of such data, a simplistic approach was adopted here. Although cross-contamination between cantaloupe and facility surfaces affected the prevalence of contaminated servings (Figure 3B), it had very low effect on the number of illnesses (Figure 3C). This suggests that the simplification of the cross-contamination process adopted in this study can be justified. However, since temperature abuse can lead to a large consumed dose, an increase in contamination prevalence, even with a low number of bacterial cells, may in the worst-case scenarios lead to a larger number of illnesses. Therefore, future research should be directed at understanding cross contamination in a facility and its effect on the risk of listeriosis.

## ACKNOWLEDGEMENTS

This work was supported through a grant (award 2019-51181-30016) from the U.S. Department of Agriculture (USDA), National Institute of Food and Agriculture (NIFA). Partial support received from a grant (award 2020-67021-32855) from the USDA NIFA.

## SUPPLEMENTAL MATERIAL

Supplemental material associated with this article can be found outline at: [URL to be completed by the publisher].

---

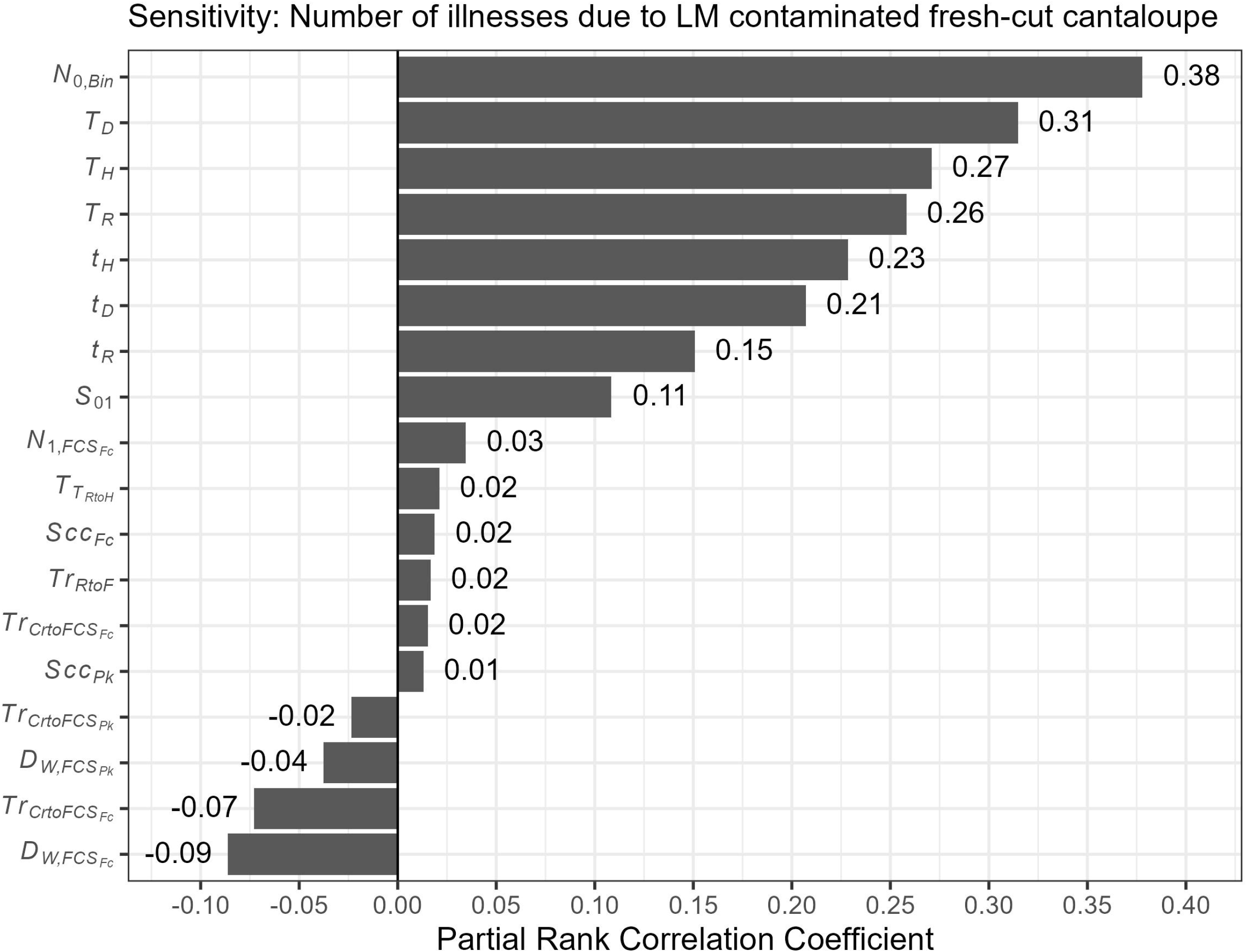

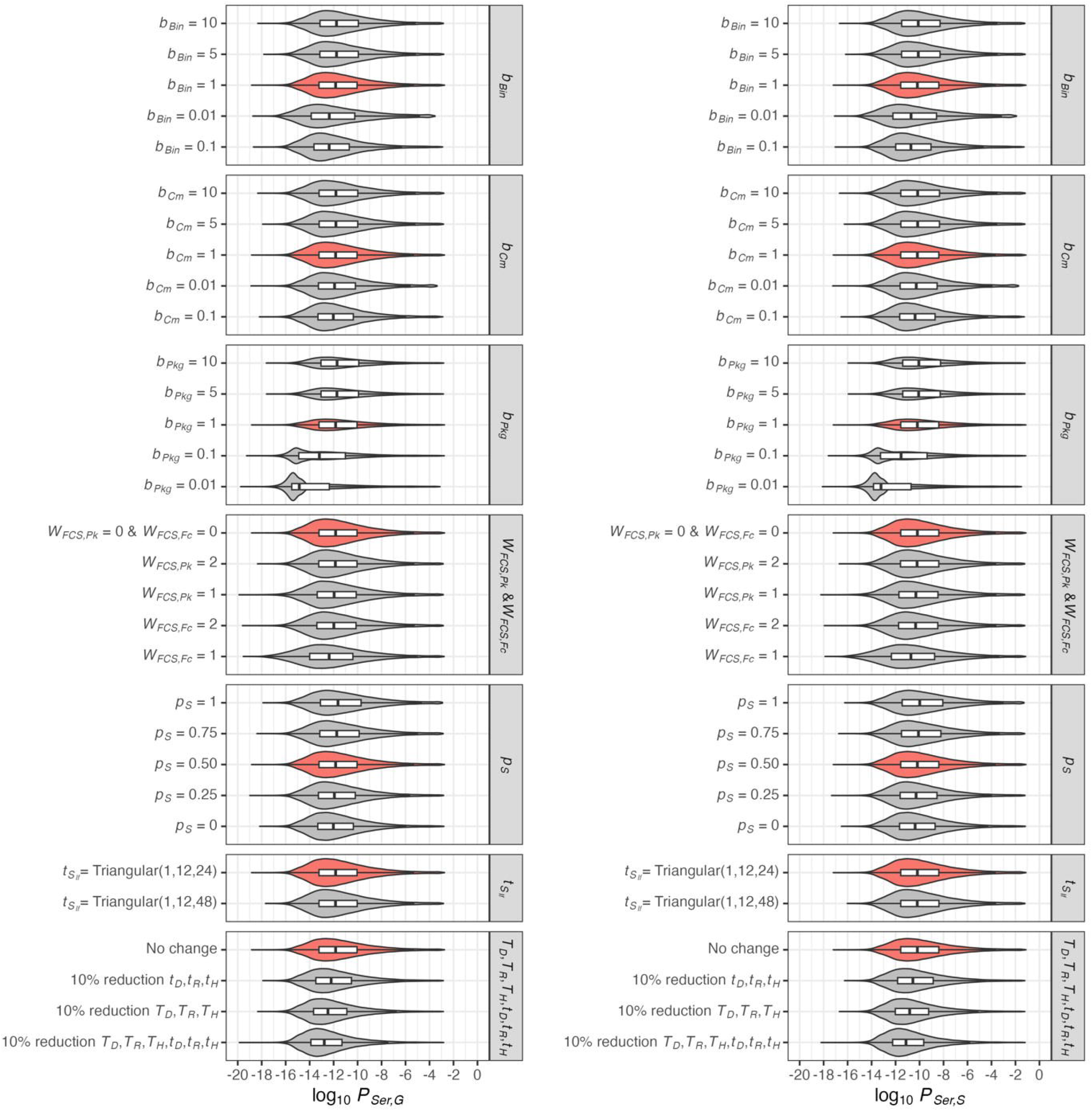

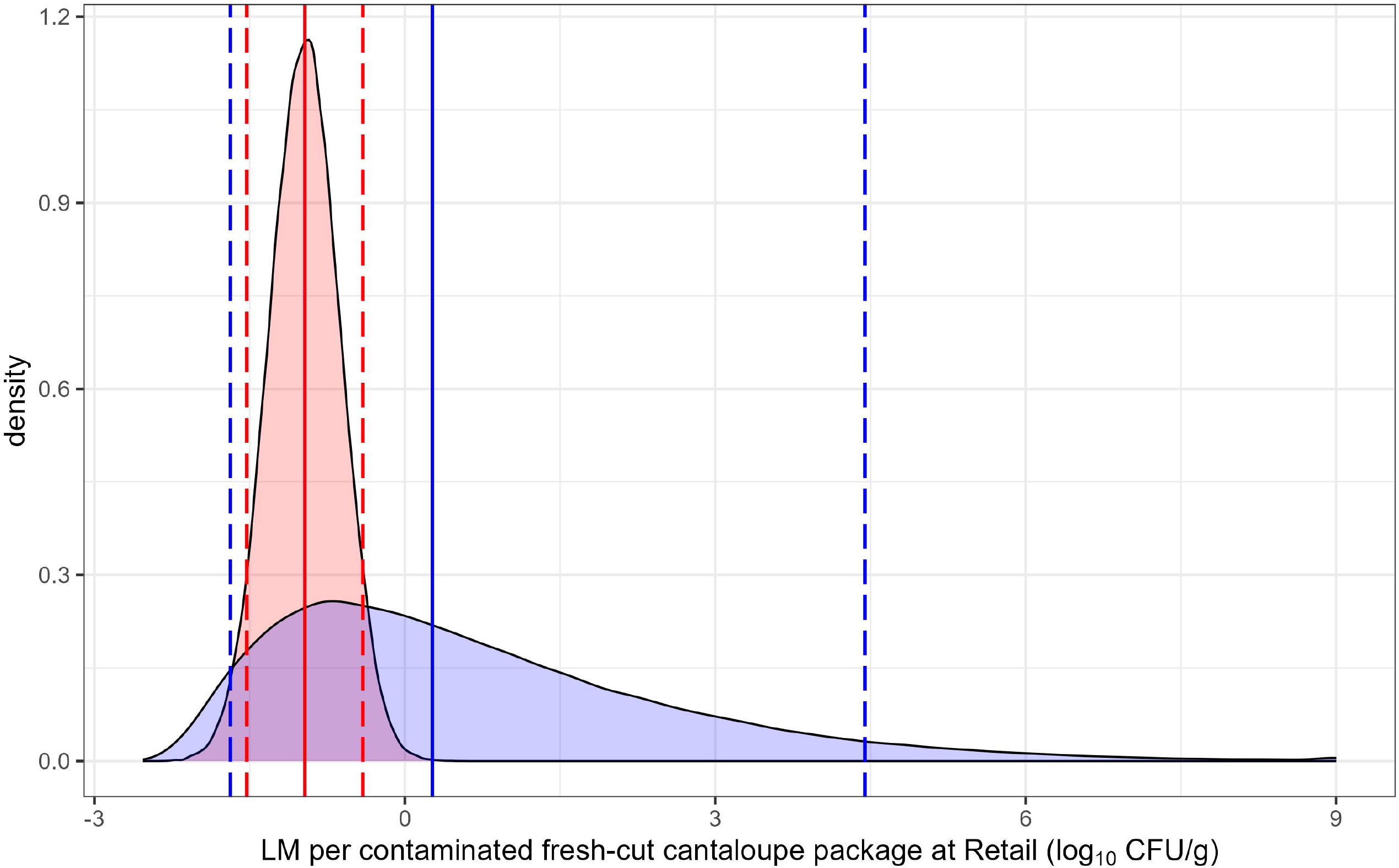

## REFERENCES

1. Bailin, D. (2013). Killer cantaloupes: ignoring the science behind food safety. https://www.sciencedirect.com/journal/journal-of-food-protection/publish/guide-for-authors/. Accessed December 20, 2024.

2. Barnett-Neefs, C., Sullivan, G., Zoellner, C., Wiedmann, M., & Ivanek, R. (2022). Using agent-based modeling to compare corrective actions for Listeria contamination in produce packinghouses. PLOS ONE, 17(3), Article e0265251. 10.1371/journal.pone.0265251.

3. Bartlett, Z., Bowman, J.P., Danyluk, M., Frankish, E., Singh, S.P., Stanley, R., & Ross, T. (2020). VM19000 Technical Report: The effective control of Listeria on whole rockmelons through alternative post-harvest treatment methods. https://www.horticulture.com.au/globalassets/laserfiche/assets/project-reports/vm19000/vm19000-final-report-complete.pdf/. Accessed December 20, 2024.

4. Buchanan, R. L., Damert, W. G., Whiting, R. C., & van Schothorst, M. (1997). Use of epidemiologic and food survey data to estimate a purposefully conservative dose-response relationship for Listeria monocytogenes levels and incidence of listeriosis. Journal of Food Protection, 60(8), 918–922. 10.4315/0362-028X-60.8.918.

5. Carstens, C. K., Salazar, J. K., & Darkoh, C. (2019). Multistate outbreaks of foodborne illness in the United States associated with fresh produce from 2010 to 2017. Frontiers in Microbiology, 10, Article 2667. 10.3389/fmicb.2019.02667.

6. Chen, Y., Dennis, S. B., Hartnett, E., Paoli, G., Pouillot, R., Ruthman, T., & Wilson, M. (2013). FDA-iRISK--a comparative risk assessment system for evaluating and ranking food-hazard pairs: case studies on microbial hazards. Journal of Food Protection, 76(3), 376–385. 10.4315/0362-028X.JFP-12-372.

7. Danyluk, M. D., Friedrich, L. M., & Schaffner, D. W. (2014). Modeling the growth of Listeria monocytogenes on cut cantaloupe, honeydew and watermelon. Food Microbiology, 38, 52–55. 10.1016/j.fm.2013.08.001.

8. De Keuckelaere, A., Jacxsens, L., Amoah, P., Medema, G., McClure, P., Jaykus, L.-A. and Uyttendaele, M. (2015). Zero risk does not exist: lessons learned from microbial risk assessment related to use of water and safety of fresh produce. Comprehensive Reviews in Food Science and Food Safety, 14, 387–410. 10.1111/1541-4337.12140.

9. Eastern Cantaloupe Growers Association. (2013). CommodityLSpecific Food Safety Guidelines for the Eastern Cantaloupes Growers Association. https://ecga-usa.org/wp-content/uploads/2017/10/ecga_commodity_specific_food_safety_guidlines_-_4.2.2013.pdf/. Accessed December 20, 2024.

10. EcoSure. U.S. Cold Temperature Survey. https://www.foodrisk.org/resources/display/21/. Accessed December 20, 2024.

11. Eifert, J. D., G. C. Sanglay, D. J. Lee, S. S. Sumner, & M. D. Pierson. (2006). Prediction of raw produce surface area from weight measurement. Journal of Food Engineering, 74, 552–556. 10.1016/j.jfoodeng.2005.02.030.

12. Fairbank, W. C., L. L. Ede, H. Johnson Jr., D. A. Luvisi, & R. A. Neja. (1987). Night picking. California Agriculture, 41, 13–16. https://californiaagriculture.org/article/110751-night-picking/. Accessed December 20, 2024.

13. Fang, T., Liu, Y., & Huang, L. (2013). Growth kinetics of Listeria monocytogenes and spoilage microorganisms in fresh-cut cantaloupe. Food Microbiology, 34(1), 174–181. 10.1016/j.fm.2012.12.005.

14. FAO/WHO. 2004. Risk assessment of Listeria monocytogenes in ready-to-eat foods. FAO/WHO Microbiological Risk Assessment Series, 4, 1–48. https://www.fao.org/fileadmin/templates/agns/pdf/jemra/mra4_en.pdf/. Accessed December 20, 2024.

15. FDA. (2015). Joint FDA/Health Canada quantitative assessment of the risk of listeriosis from soft-ripened cheese consumption in the United States and Canada: Report. https://www.fda.gov/food/risk-and-safety-assessments-food/joint-fda-health-canada-quantitative-assessment-risk-listeriosis-soft-ripened-cheese-consumption/. Accessed December 20, 2024.

16. Gallagher, D., Pouillot, R., Hoelzer, K., Tang, J., Dennis, S. B., & Kause, J. R. (2016). Listeria monocytogenes in retail delicatessens: an interagency risk assessment-risk mitigations. Journal of Food Protection, 79(7), 1076–1088. 10.4315/0362-028X.JFP-15-336.

17. Harrand, A. S., Guariglia-Oropeza, V., Skeens, J., Kent, D., & Wiedmann, M. (2021). Nature versus nurture: assessing the impact of strain diversity and pregrowth conditions on Salmonella enterica, Escherichia coli, and Listeria species growth and survival on selected produce items. Applied and Environmental Microbiology, 87(6), Article e01925–20. 10.1128/AEM.01925-20.

18. Hoelzer, K., Pouillot, R., Gallagher, D., Silverman, M. B., Kause, J., & Dennis, S. (2012). Estimation of Listeria monocytogenes transfer coefficients and efficacy of bacterial removal through cleaning and sanitation. International Journal of Food Microbiology, 157(2), 267–277. 10.1016/j.ijfoodmicro.2012.05.019.

19. Latorre, A. A., Pradhan, A. K., Van Kessel, J. A., Karns, J. S., Boor, K. J., Rice, D. H., Mangione, K. J., Gröhn, Y. T., & Schukken, Y. H. (2011). Quantitative risk assessment of listeriosis due to consumption of raw milk. Journal of Food Protection, 74(8), 1268–1281. 10.4315/0362-028X.JFP-10-554.

20. Liao, J., Guo, X., Weller, D. L., Pollak, S., Buckley, D. H., Wiedmann, M., & Cordero, O. X. (2021). Nationwide genomic atlas of soil-dwelling Listeria reveals effects of selection and population ecology on pangenome evolution. Nature Microbiology, 6(8), 1021–1030. 10.1038/s41564-021-00935-7.

21. McCollum, J. T., Cronquist, A. B., Silk, B. J., Jackson, K. A., O’Connor, K. A., Cosgrove, S., Gossack, J. P., Parachini, S. S., Jain, N. S., Ettestad, P., Ibraheem, M., Cantu, V., Joshi, M., DuVernoy, T., Fogg, N. W., Jr, Gorny, J.R., Mogen, K. M., Spires, C., Teitell, P., Joseph, L. A., Tarr, C. L., Imanishi, M., Neil, K. P., Tauxe, R. V., & Mahon, B. E. (2013). Multistate outbreak of listeriosis associated with cantaloupe. The New England Journal of Medicine, 369(10), 944– 953. 10.1056/NEJMoa1215837.

22. Mokhtari, A., Pang, H., Santillana Farakos, S., McKenna, C., Crowley, C., Cranford, V., Bowen, A., Phillips, S., Madad, A., Obenhuber, D., & Van Doren, J. M. (2023). Leveraging risk assessment for foodborne outbreak investigations: The Quantitative risk assessment-epidemic curve prediction model. Risk Analysis: An official publication of the Society for Risk Analysis, 43(2), 324–338. 10.1111/risa.13896.

23. Nauta, M. J. (2008). The modular process risk model (MPRM): A structured approach to food chain exposure assessment, 99–136. D.W. Schaffner (ed.), Microbial Risk Analysis of Foods, ASM Press, Washington, D.C. 10.1128/9781555815752.ch4.

24. Nauta M. J. (2005). Microbiological risk assessment models for partitioning and mixing during food handling. International Journal of Food Microbiology, 100(1-3), 311–322. 10.1016/j.ijfoodmicro.2004.10.027.

25. NSW Department of Primary Industries. (2018). Listeria Outbreak Investigation - Summary Report for the Melon Industry, October 2018. https://www.foodauthority.nsw.gov.au/sites/default/files/_Documents/foodsafetyandyou/listeria_outbreak_investigation.pdf/. Accessed December 20, 2024.

26. Pang, H., Lambertini, E., Buchanan, R. L., Schaffner, D. W., & Pradhan, A. K. (2017). Quantitative microbial risk assessment for Escherichia coli O157:H7 in fresh-cut lettuce. Journal of Food Protection, 80(2), 302–311. 10.4315/0362-028X.JFP-16-246.

27. Pouillot, R., Chen, Y., & Hoelzer, K. (2015). Modeling number of bacteria per food unit in comparison to bacterial concentration in quantitative risk assessment: impact on risk estimates. Food Microbiology, 45(Pt B), 245–253. 10.1016/j.fm.2014.05.008.

28. Pouillot, R., Lubran, M. B., Cates, S. C., & Dennis, S. (2010). Estimating parametric distributions of storage time and temperature of ready-to-eat foods for U.S. households. Journal of Food Protection, 73(2), 312–321. 10.4315/0362-028x-73.2.312.

29. R Core Team. (2021). R: A language and environment for statistical computing. R Foundation for Statistical Computing. Vienna, Austria. https://www.r-project.org/. Accessed December 20, 2024.

30. Salazar, J. K., Sahu, S. N., Hildebrandt, I. M., Zhang, L., Qi, Y., Liggans, G., Datta, A. R., & Tortorello, M. L. (2017). Growth kinetics of Listeria monocytogenes in cut produce. Journal of Food Protection, 80(8), 1328–1336. 10.4315/0362-028X.JFP-16-516.

31. Singh, S. P. (2019). Melon food safety: A best practice guide for rockmelons and specialty melons. https://fpsc-anz.com/wp-content/uploads/2019/09/Melon-food-safety-best-practice-guide.pdf/. Accessed December 20, 2024.

32. Stevenson, M., & E. Sergeant. (2022). epiR: Tools for the Analysis of Epidemiological Data. https://cran.r-project.org/web/packages/epiR/index.html/. Accessed December 20, 2024.

33. Sullivan, G., Zoellner, C., Wiedmann, M., & Ivanek, R. (2021). In silico models for design and optimization of science-based Listeria environmental monitoring programs in fresh-cut produce facilities. Applied and Environmental Microbiology, 87(21), Article e0079921. 10.1128/AEM.00799-21.

34. Swaminathan, B., & Gerner-Smidt, P. (2007). The epidemiology of human listeriosis. Microbes and Infection, 9(10), 1236–1243. 10.1016/j.micinf.2007.05.011.

35. Topalcengiz, Z., J. Saha, L. M. Friedrich, L. D. Goodridge, and M. D. Danyluk. (2022). Fate of Escherichia coli O157:H7, Salmonella, and Listeria monocytogenes on whole cantaloupe and watermelon surfaces during storage. SSRN: 10.2139/ssrn.4471803.

36. Torres, A., Langenhoven, P., & Behe, B. K. (2020). Characterizing the U.S. melon market. HortScience, 55(6), 795–803. 10.21273/HORTSCI14859-20/.

37. Townsend, A., Strawn, L. K., Chapman, B. J., & Dunn, L. L. (2021). A systematic review of Listeria species and Listeria monocytogenes prevalence, persistence, and diversity throughout the fresh produce supply chain. Foods (Basel, Switzerland), 10(6), 1427. 10.3390/foods10061427.

38. U.S. Census Bureau. Annual estimates of the resident population for the United States, states, and the District of Columbia: April 1, 2010 to July 1, 2020 (NST-EST2020). https://www.census.gov/programs-surveys/popest/technical-documentation/research/evaluation-estimates/2020-evaluation-estimates/2010s-state-total.html/. Accessed December 20, 2024.

39. U.S. Department of Agriculture. Melons, cantaloupe, raw - FoodData Central - USDA.https://fdc.nal.usda.gov/. Accessed December 20, 2024.

40. U.S. FDA. (2013). National commodity-specific food safety guidelines for cantaloupes and netted melons, version 1.1. https://www.fda.gov/files/food/published/Commodity-Specific-Food-Safety-Guidelines-for-Cantaloupes-and-Netted-Melons-%28PDF%29.pdf/. Accessed December 20, 2024.

41. Ukuku, D. O., Huang, L., & Sommers, C. (2015). Efficacy of sanitizer treatments on survival and growth parameters of Escherichia coli O157:H7, Salmonella, and Listeria monocytogenes on fresh-cut pieces of cantaloupe during storage. Journal of Food Protection, 78(7), 1288–1295. 10.4315/0362-028X.JFP-14-233.

42. Ukuku, D. O., Olanya, M., Geveke, D. J., & Sommers, C. H. (2012). Effect of native microflora, waiting period, and storage temperature on Listeria monocytogenes serovars transferred from cantaloupe rind to fresh-cut pieces during preparation. Journal of Food Protection, 75(11), 1912– 1919. 10.4315/0362-028X.JFP-12-191.

43. United States Department of Agriculture Economic Research Service. Food Availability (Per Capita) Data System. https://www.ers.usda.gov/data-products/food-availability-per-capita-data-system/. Accessed December 20, 2024.

44. Walsh, K. A., Bennett, S. D., Mahovic, M., & Gould, L. H. (2014). Outbreaks associated with cantaloupe, watermelon, and honeydew in the United States, 1973-2011. Foodborne Pathogens and Disease, 11(12), 945–952. 10.1089/fpd.2014.1812.

45. Zhang, G., Chen, Y., Hu, L., Melka, D., Wang, H., Laasri, A., Brown, E. W., Strain, E., Allard, M., Bunning, V. K., Parish, M., Musser, S. M., & Hammack, T. S. (2018). Survey of foodborne pathogens, aerobic plate counts, total coliform counts, and Escherichia coli counts in leafy greens, sprouts, and melons marketed in the United States. Journal of Food Protection, 81(3), 400–411. 10.4315/0362-028X.JFP-17-253.

46. Zoellner, C., Wiedmann, M., & Ivanek, R. (2019). An assessment of listeriosis risk associated with a contaminated production lot of frozen vegetables consumed under alternative consumer handling scenarios. Journal of Food Protection, 82(12), 2174–2193. 10.4315/0362-028X.JFP-19-092.

