## Supplemental Material for "Farm-to-consumer quantitative microbial risk assessment model for *Listeria monocytogenes* on fresh-cut cantaloupe"

Postal address: 602 Tower Rd, Ithaca, NY 14853

Supplemental Table 1. Summary of results from scenario analysis: $p_S$

| | General Population | | | | Susceptible Population | | | |
|---|---|---|---|---|---|---|---|---|
| $p_S$ | Median | 5% | 95% | 99% | Median | $5^{th}$ | $95^{th}$ | $99^{th}$ |
| 0.0 | $9.5 \times 10^{-13}$ | $1.6 \times 10^{-15}$ | $6.9 \times 10^{-8}$ | $4.5 \times 10^{-5}$ | $4.2 \times 10^{-11}$ | $7.2 \times 10^{-14}$ | $3.1 \times 10^{-6}$ | $1.9 \times 10^{-3}$ |
| 0.25 | $1.2 \times 10^{-12}$ | $1.7 \times 10^{-15}$ | $1.3 \times 10^{-7}$ | $7.8 \times 10^{-5}$ | $5.2 \times 10^{-11}$ | $7.6 \times 10^{-14}$ | $5.8 \times 10^{-6}$ | $3.4 \times 10^{-3}$ |
| 0.5 (baseline) | $1.4 \times 10^{-12}$ | $1.8 \times 10^{-15}$ | $2.4 \times 10^{-7}$ | $1.1 \times 10^{-4}$ | $6.4 \times 10^{-11}$ | $7.9 \times 10^{-14}$ | $1.1 \times 10^{-5}$ | $4.8 \times 10^{-3}$ |
| 0.75 | $1.8 \times 10^{-12}$ | $1.9 \times 10^{-15}$ | $4.5 \times 10^{-7}$ | $1.6 \times 10^{-4}$ | $7.9 \times 10^{-11}$ | $8.5 \times 10^{-14}$ | $2.0 \times 10^{-5}$ | $7.1 \times 10^{-3}$ |
| 1.0 | $2.3 \times 10^{-12}$ | $2.1 \times 10^{-15}$ | $7.5 \times 10^{-7}$ | $2.3 \times 10^{-4}$ | $1.0 \times 10^{-10}$ | $9.2 \times 10^{-14}$ | $3.3 \times 10^{-5}$ | $9.9 \times 10^{-3}$ |

| | Number of illnesses | | | | Probability of ≥1 illness | Number of deaths | | | | Probability of ≥1 death |
|---|---|---|---|---|---|---|---|---|---|---|
| $p_S$ | Median | 5% | 95% | 99% | | Median | 5% | 95% | 99% | |
| 0.0 | 0 | 0 | 309 | 193,695 | 20.8% | 0 | 0 | 75 | 48,510 | 15.7% |
| 0.25 | 0 | 0 | 578 | 338,204 | 22.5% | 0 | 0 | 142 | 85,002 | 17.3% |
| 0.5 (baseline) | 0 | 0 | 1,070 | 475,436 | 24.1% | 0 | 0 | 264 | 119,639 | 18.8% |
| 0.75 | 0 | 0 | 1,973 | 696,698 | 25.7% | 0 | 0 | 490 | 174,946 | 20.3% |
| 1.0 | 0 | 0 | 3,287 | 986,276 | 27.4% | 0 | 0 | 824 | 248,563 | 21.8% |

Supplemental Table 2. Summary of results from scenario analysis: $b_{Bin}$

| | General Population | | | | Susceptible Population | | | |
|---|---|---|---|---|---|---|---|---|
| $b_{Bin}$ | Median | 5% | 95% | 99% | Median | $5^{th}$ | $95^{th}$ | $99^{th}$ |
| 0.01 | $4.7 \times 10^{-13}$ | $3.4 \times 10^{-16}$ | $5.7 \times 10^{-7}$ | $6.4 \times 10^{-5}$ | $2.1 \times 10^{-11}$ | $1.5 \times 10^{-14}$ | $2.5 \times 10^{-5}$ | $2.7 \times 10^{-3}$ |
| 0.1 | $4.3 \times 10^{-13}$ | $8.3 \times 10^{-16}$ | $6.1 \times 10^{-8}$ | $3.9 \times 10^{-5}$ | $1.9 \times 10^{-11}$ | $3.7 \times 10^{-14}$ | $2.7 \times 10^{-6}$ | $1.7 \times 10^{-3}$ |
| 1 (baseline) | $1.4 \times 10^{-12}$ | $1.8 \times 10^{-15}$ | $2.4 \times 10^{-7}$ | $1.1 \times 10^{-4}$ | $6.4 \times 10^{-11}$ | $7.9 \times 10^{-14}$ | $1.1 \times 10^{-5}$ | $4.8 \times 10^{-3}$ |
| 5 | $1.7 \times 10^{-12}$ | $2.0 \times 10^{-15}$ | $2.9 \times 10^{-7}$ | $1.3 \times 10^{-4}$ | $7.8 \times 10^{-11}$ | $9.1 \times 10^{-14}$ | $1.3 \times 10^{-5}$ | $5.8 \times 10^{-3}$ |
| 10 | $1.8 \times 10^{-12}$ | $2.1 \times 10^{-15}$ | $3.1 \times 10^{-7}$ | $1.4 \times 10^{-4}$ | $7.9 \times 10^{-11}$ | $9.2 \times 10^{-14}$ | $1.4 \times 10^{-5}$ | $5.9 \times 10^{-3}$ |

| | Number of illnesses | | | | Probability of ≥1 illness | Number of deaths | | | | Probability of ≥1 death |
|---|---|---|---|---|---|---|---|---|---|---|
| $b_{Bin}$ | Median | 5% | 95% | 99% | | Median | 5% | 95% | 99% | |
| 0.01 | 0 | 0 | 2,473 | 267,301 | 22.2% | 0 | 0 | 618 | 67,005 | 17.9% |
| 0.1 | 0 | 0 | 268 | 167,310 | 18.1% | 0 | 0 | 67 | 41,543 | 13.8% |
| 1 (baseline) | 0 | 0 | 1,070 | 475,436 | 24.1% | 0 | 0 | 264 | 119,639 | 18.8% |
| 5 | 0 | 0 | 1,314 | 574,034 | 25.1% | 0 | 0 | 324 | 143,338 | 19.6% |
| 10 | 0 | 0 | 1,353 | 578,345 | 25.2% | 0 | 0 | 3331 | 146,415 | 19.7% |

Supplemental Table 3. Summary of results from scenario analysis: $b_{Cm}$

| | General Population | | | | Susceptible Population | | | |
|---|---|---|---|---|---|---|---|---|
| $b_{Cm}$ | Median | 5% | 95% | 99% | Median | $5^{th}$ | $95^{th}$ | $99^{th}$ |
| 0.01 | $1.2 \times 10^{-12}$ | $1.7 \times 10^{-15}$ | $1.8 \times 10^{-7}$ | $8.2 \times 10^{-5}$ | $5.4 \times 10^{-11}$ | $7.5 \times 10^{-14}$ | $8.0 \times 10^{-6}$ | $3.5 \times 10^{-3}$ |
| 0.1 | $9.3 \times 10^{-13}$ | $1.7 \times 10^{-15}$ | $8.1 \times 10^{-8}$ | $5.6 \times 10^{-5}$ | $4.2 \times 10^{-11}$ | $7.6 \times 10^{-14}$ | $3.6 \times 10^{-6}$ | $2.4 \times 10^{-3}$ |
| 1 (baseline) | $1.4 \times 10^{-12}$ | $1.8 \times 10^{-15}$ | $2.4 \times 10^{-7}$ | $1.1 \times 10^{-4}$ | $6.4 \times 10^{-11}$ | $7.9 \times 10^{-14}$ | $1.1 \times 10^{-5}$ | $4.8 \times 10^{-3}$ |
| 5 | $1.5 \times 10^{-12}$ | $1.8 \times 10^{-15}$ | $3.0 \times 10^{-7}$ | $1.3 \times 10^{-4}$ | $6.9 \times 10^{-11}$ | $8.1 \times 10^{-14}$ | $1.3 \times 10^{-5}$ | $5.5 \times 10^{-3}$ |
| 10 | $1.6 \times 10^{-12}$ | $1.8 \times 10^{-15}$ | $3.0 \times 10^{-7}$ | $1.3 \times 10^{-4}$ | $6.9 \times 10^{-11}$ | $8.1 \times 10^{-14}$ | $1.4 \times 10^{-5}$ | $5.6 \times 10^{-3}$ |

| | Number of illnesses | | | | Probability of ≥1 illness | Number of deaths | | | | Probability of ≥1 death |
|---|---|---|---|---|---|---|---|---|---|---|
| $b_{Cm}$ | Median | 5% | 95% | 99% | | Median | 5% | 95% | 99% | |
| 0.01 | 0 | 0 | 785 | 344,762 | 22.8% | 0 | 0 | 194 | 87,017 | 17.7% |
| 0.1 | 0 | 0 | 354 | 237,089 | 20.7% | 0 | 0 | 87 | 59,093 | 15.7% |
| 1 (baseline) | 0 | 0 | 1,070 | 475,436 | 24.1% | 0 | 0 | 264 | 119,639 | 18.8% |
| 5 | 0 | 0 | 1,303 | 543,627 | 24.7% | 0 | 0 | 321 | 133,837 | 19.2% |
| 10 | 0 | 0 | 1,326 | 552,936 | 24.7% | 0 | 0 | 325 | 138,770 | 19.3% |

Supplemental Table 4. Summary of results from scenario analysis: $b_{Pkg}$

| | General Population | | | | Susceptible Population | | | |
|---|---|---|---|---|---|---|---|---|
| $b_{Pkg}$ | Median | 5% | 95% | 99% | Median | $5^{th}$ | $95^{th}$ | $99^{th}$ |
| 0.01 | $1.4 \times 10^{-15}$ | $5.7 \times 10^{-17}$ | $4.4 \times 10^{-8}$ | $5.5 \times 10^{-5}$ | $6.1 \times 10^{-14}$ | $2.6 \times 10^{-15}$ | $2.0 \times 10^{-6}$ | $2.4 \times 10^{-3}$ |
| 0.1 | $6.5 \times 10^{-14}$ | $1.7 \times 10^{-16}$ | $1.3 \times 10^{-7}$ | $1.1 \times 10^{-4}$ | $2.9 \times 10^{-12}$ | $7.6 \times 10^{-15}$ | $5.8 \times 10^{-6}$ | $4.6 \times 10^{-3}$ |
| 1 (baseline) | $1.4 \times 10^{-12}$ | $1.8 \times 10^{-15}$ | $2.4 \times 10^{-7}$ | $1.1 \times 10^{-4}$ | $6.4 \times 10^{-11}$ | $7.9 \times 10^{-14}$ | $1.1 \times 10^{-5}$ | $4.8 \times 10^{-3}$ |
| 5 | $1.9 \times 10^{-12}$ | $3.5 \times 10^{-15}$ | $2.9 \times 10^{-7}$ | $1.1 \times 10^{-4}$ | $8.4 \times 10^{-11}$ | $1.6 \times 10^{-13}$ | $1.3 \times 10^{-5}$ | $4.8 \times 10^{-3}$ |
| 10 | $1.9 \times 10^{-12}$ | $3.7 \times 10^{-15}$ | $3.0 \times 10^{-7}$ | $1.1 \times 10^{-4}$ | $8.7 \times 10^{-11}$ | $1.7 \times 10^{-13}$ | $1.3 \times 10^{-5}$ | $4.8 \times 10^{-3}$ |

| | Number of illnesses | | | | Probability of ≥1 illness | Number of deaths | | | | Probability of ≥1 death |
|---|---|---|---|---|---|---|---|---|---|---|
| $b_{Pkg}$ | Median | 5% | 95% | 99% | | Median | 5% | 95% | 99% | |
| 0.01 | 0 | 0 | 196 | 231,652 | 11.5% | 0 | 0 | 49 | 58,697 | 9.4% |
| 0.1 | 0 | 0 | 567 | 450,973 | 16.6% | 0 | 0 | 142 | 114,029 | 13.1% |
| 1 (baseline) | 0 | 0 | 1,070 | 475,436 | 24.1% | 0 | 0 | 264 | 119,639 | 18.8% |
| 5 | 0 | 0 | 1,265 | 475,436 | 25.1% | 0 | 0 | 315 | 119,639 | 19.6% |
| 10 | 0 | 0 | 1,298 | 475,436 | 25.3% | 0 | 0 | 325 | 119,639 | 19.7% |

Supplemental Table 5. Summary of results from scenario analysis: Sanitation schedule for FCS at Packinghouse and Fresh-cut facility

| | General Population | | | | Susceptible Population | | | |
|---|---|---|---|---|---|---|---|---|
| $W_{FCS,Fc}, W_{FCS,Pk}$ | Median | 5% | 95% | 99% | Median | 5th | 95th | 99th |
| $W_{FCS,Fc} = 0, W_{FCS,Pk} = 0$ (baseline) | $1.4 \times 10^{-12}$ | $1.8 \times 10^{-15}$ | $2.4 \times 10^{-7}$ | $1.1 \times 10^{-4}$ | $6.4 \times 10^{-11}$ | $7.9 \times 10^{-14}$ | $1.1 \times 10^{-5}$ | $4.8 \times 10^{-3}$ |
| $W_{FCS,Pk} = 1$ | $1.1 \times 10^{-12}$ | $9.4 \times 10^{-16}$ | $2.2 \times 10^{-7}$ | $1.1 \times 10^{-4}$ | $4.9 \times 10^{-11}$ | $4.2 \times 10^{-14}$ | $9.6 \times 10^{-6}$ | $4.6 \times 10^{-3}$ |
| $W_{FCS,Pk} = 2$ | $1.4 \times 10^{-12}$ | $1.6 \times 10^{-15}$ | $2.4 \times 10^{-7}$ | $1.1 \times 10^{-4}$ | $6.0 \times 10^{-11}$ | $7.3 \times 10^{-14}$ | $1.1 \times 10^{-5}$ | $4.7 \times 10^{-3}$ |
| $W_{FCS,Fc} = 1$ | $4.1 \times 10^{-13}$ | $1.0 \times 10^{-15}$ | $1.5 \times 10^{-7}$ | $9.2 \times 10^{-5}$ | $1.8 \times 10^{-11}$ | $4.5 \times 10^{-15}$ | $6.7 \times 10^{-6}$ | $4.0 \times 10^{-3}$ |
| $W_{FCS,Fc} = 2$ | $1.0 \times 10^{-12}$ | $1.0 \times 10^{-15}$ | $2.1 \times 10^{-7}$ | $1.0 \times 10^{-4}$ | $4.6 \times 10^{-11}$ | $4.5 \times 10^{-14}$ | $9.3 \times 10^{-6}$ | $4.4 \times 10^{-3}$ |

| | Number of illnesses | | | | Probability of ≥1 illness | Number of deaths | | | | Probability of ≥1 death |
|---|---|---|---|---|---|---|---|---|---|---|
| $W_{FCS,Fc}, W_{FCS,Pk}$ | Median | 5% | 95% | 99% | | Median | 5% | 95% | 99% | |
| $W_{FCS,Fc} = 0, W_{FCS,Pk} = 0$ (baseline) | 0 | 0 | 1,070 | 475,436 | 24.1% | 0 | 0 | 264 | 119,639 | 18.8% |
| $W_{FCS,Pk} = 1$ | 0 | 0 | 945 | 449,200 | 23.3% | 0 | 0 | 236 | 113,573 | 18.2% |
| $W_{FCS,Pk} = 2$ | 0 | 0 | 1,037 | 466,189 | 23.9% | 0 | 0 | 259 | 118,518 | 18.7% |
| $W_{FCS,Fc} = 1$ | 0 | 0 | 665 | 390,565 | 20.9% | 0 | 0 | 164 | 97,127 | 16.5% |
| $W_{FCS,Fc} = 2$ | 0 | 0 | 917 | 432,115 | 23.1% | 0 | 0 | 230 | 107,783 | 18.0% |

Supplemental Table 6. Summary of results from scenario analysis: Time and temperature at Distribution, Retail storage/display, and Home storage

| | General Population | | | | Susceptible Population | | | |
|---|---|---|---|---|---|---|---|---|
| $t_D, t_R, t_H$ $T_D, T_R, T_H$ | Median | 5% | 95% | 99% | Median | 5th | 95th | 99th |
| (baseline) | $1.4 \times 10^{-12}$ | $1.8 \times 10^{-15}$ | $2.4 \times 10^{-7}$ | $1.1 \times 10^{-4}$ | $6.4 \times 10^{-11}$ | $7.9 \times 10^{-14}$ | $1.1 \times 10^{-5}$ | $4.8 \times 10^{-3}$ |
| -10%_$t_D, t_R, t_H$ | $6.2 \times 10^{-13}$ | $1.2 \times 10^{-15}$ | $5.1 \times 10^{-8}$ | $3.1 \times 10^{-5}$ | $2.8 \times 10^{-11}$ | $5.5 \times 10^{-14}$ | $2.3 \times 10^{-6}$ | $1.4 \times 10^{-3}$ |
| -10%_$T_D, T_R, T_H$ | $3.3 \times 10^{-13}$ | $9.8 \times 10^{-16}$ | $1.6 \times 10^{-8}$ | $8.3 \times 10^{-6}$ | $1.5 \times 10^{-11}$ | $4.4 \times 10^{-14}$ | $7.3 \times 10^{-7}$ | $3.7 \times 10^{-4}$ |
| -10%_$t_D, t_R, t_H,$ $T_D, T_R, T_H$ | $1.7 \times 10^{-13}$ | $7.4 \times 10^{-16}$ | $5.1 \times 10^{-9}$ | $1.7 \times 10^{-6}$ | $7.4 \times 10^{-12}$ | $3.3 \times 10^{-14}$ | $2.3 \times 10^{-7}$ | $7.7 \times 10^{-5}$ |

| | Number of illnesses | | | | Probability of ≥1 illness | Number of deaths | | | | Probability of ≥1 death |
|---|---|---|---|---|---|---|---|---|---|---|
| $t_D, t_R, t_H$ $T_D, T_R, T_H$ | Median | 5% | 95% | 99% | | Median | 5% | 95% | 99% | |

| | | | | | | | | | | |
|---|---|---|---|---|---|---|---|---|---|---|
| (baseline) | 0 | 0 | 1,070 | 475,436 | 24.1% | 0 | 0 | 264 | 119,639 | 18.8% |
| -10%_$t_D, t_R, t_H$ | 0 | 0 | 224 | 134,673 | 19.4% | 0 | 0 | 56 | 33,668 | 14.6% |
| -10%_$T_D, T_R, T_H$ | 0 | 0 | 72 | 1,075 | 15.9% | 0 | 0 | 18 | 9,017 | 11.7% |
| -10%_$t_D, t_R, t_H,$ $T_D, T_R, T_H$ | 0 | 0 | 22 | 7,361 | 12.7% | 0 | 0 | 6 | 1,888 | 9.2% |

Supplemental Table 7. Summary of results from scenario analysis: Extended storage time at the intermediate facility

| | General Population | | | | Susceptible Population | | | |
|---|---|---|---|---|---|---|---|---|
| $t_{S_{If}}$ | Median | 5% | 95% | 99% | Median | 5th | 95th | 99th |
| (baseline) | 1.4 x $10^{-12}$ | 1.8 x $10^{-15}$ | 2.4 x $10^{-7}$ | 1.1 x $10^{-4}$ | 6.4 x $10^{-11}$ | 7.9 x $10^{-14}$ | 1.1 x $10^{-5}$ | 4.8 x $10^{-3}$ |
| Ext-$t_{S_{If}}$ | 1.4 x $10^{-12}$ | 1.8 x $10^{-15}$ | 2.4 x $10^{-7}$ | 1.1 x $10^{-4}$ | 6.4 x $10^{-11}$ | 8.0 x $10^{-14}$ | 1.1 x $10^{-5}$ | 4.9 x $10^{-3}$ |

| | Number of illnesses | | | | Probability of ≥1 illness | Number of deaths | | | | Probability of ≥1 death |
|---|---|---|---|---|---|---|---|---|---|---|
| $t_{S_{If}}$ | Median | 5% | 95% | 99% | | Median | 5% | 95% | 99% | |
| (baseline) | 0 | 0 | 1,070 | 475,436 | 24.1% | 0 | 0 | 264 | 119,639 | 18.8% |
| Ext-$t_{S_{If}}$ | 0 | 0 | 1,081 | 478,666 | 24.1% | 0 | 0 | 267 | 120,458 | 18.8% |

Supplemental Table 8. Summary of results from additional scenario analysis: Temperature at Distribution, Retail storage/display, and Home storage

| | General Population | | | | Susceptible Population | | | |
|---|---|---|---|---|---|---|---|---|
| $T_D, T_R, T_H$ | Median | 5% | 95% | 99% | Median | 5th | 95th | 99th |
| (baseline) | 1.4 x $10^{-12}$ | 1.8 x $10^{-15}$ | 2.4 x $10^{-7}$ | 1.1 x $10^{-4}$ | 6.4 x $10^{-11}$ | 7.9 x $10^{-14}$ | 1.1 x $10^{-5}$ | 4.8 x $10^{-3}$ |
| -10%_$T_D$ | 7.3 x $10^{-13}$ | 1.3 x $10^{-15}$ | 5.6 x $10^{-8}$ | 5.6 x $10^{-5}$ | 3.3 x $10^{-11}$ | 5.8 x $10^{-14}$ | 3.8 x $10^{-6}$ | 2.5 x $10^{-3}$ |
| -10%_$T_R$ | 9.2 x $10^{-13}$ | 1.5 x $10^{-15}$ | 1.0 x $10^{-7}$ | 6.2 x $10^{-5}$ | 4.1 x $10^{-11}$ | 6.8 x $10^{-14}$ | 4.5 x $10^{-6}$ | 2.7 x $10^{-3}$ |
| -10%_$T_H$ | 9.9 x $10^{-13}$ | 1.5 x $10^{-15}$ | 1.0 x $10^{-7}$ | 5.6 x $10^{-5}$ | 4.4 x $10^{-11}$ | 6.8 x $10^{-14}$ | 4.7 x $10^{-6}$ | 2.6 x $10^{-3}$ |
| TruncateMax_$T_D$ | 4.6 x $10^{-13}$ | 1.2 x $10^{-15}$ | 4.1 x $10^{-8}$ | 3.0 x $10^{-5}$ | 2.1 x $10^{-11}$ | 5.3 x $10^{-14}$ | 1.8 x $10^{-6}$ | 1.3 x $10^{-3}$ |
| TruncateMax_$T_R$ | 6.6 x $10^{-13}$ | 1.4 x $10^{-15}$ | 4.5 x $10^{-8}$ | 2.9 x $10^{-5}$ | 3.0 x $10^{-11}$ | 6.2 x $10^{-14}$ | 2.0 x $10^{-6}$ | 1.3 x $10^{-3}$ |
| TruncateMax_$T_H$ | 1.1 x $10^{-12}$ | 1.7 x $10^{-15}$ | 7.9 x $10^{-8}$ | 3.3 x $10^{-5}$ | 4.8 x $10^{-11}$ | 7.4 x $10^{-14}$ | 3.5 x $10^{-6}$ | 1.5 x $10^{-3}$ |
| ShiftMean_$T_D$ | 8.3 x $10^{-13}$ | 1.2 x $10^{-15}$ | 1.4 x $10^{-7}$ | 8.5 x $10^{-5}$ | 3.7 x $10^{-11}$ | 5.5 x $10^{-14}$ | 6.1 x $10^{-6}$ | 3.7 x $10^{-3}$ |
| ShiftMean_$T_R$ | 6.3 x $10^{-13}$ | 1.2 x $10^{-15}$ | 6.6 x $10^{-8}$ | 4.5 x $10^{-5}$ | 2.8 x $10^{-11}$ | 5.3 x $10^{-14}$ | 3.0 x $10^{-6}$ | 2.0 x $10^{-3}$ |
| ShiftMean_$T_H$ | 6.9 x $10^{-13}$ | 1.2 x $10^{-15}$ | 7.1 x $10^{-8}$ | 4.5 x $10^{-5}$ | 3.1 x $10^{-11}$ | 5.5 x $10^{-14}$ | 3.2 x $10^{-6}$ | 2.0 x $10^{-3}$ |
| TruncateMax_$T_D$and ShiftMean_$T_D$ | 2.9 x $10^{-13}$ | 8.7 x $10^{-16}$ | 2.6 x $10^{-8}$ | 2.0 x $10^{-5}$ | 1.3 x $10^{-11}$ | 3.9 x $10^{-14}$ | 1.2 x $10^{-6}$ | 8.8 x $10^{-4}$ |
| TruncateMax_$T_R$and | 4.4 x $10^{-13}$ | 1.1 x $10^{-15}$ | 3.0 x $10^{-8}$ | 2.0 x $10^{-5}$ | 2.0 x $10^{-11}$ | 4.9 x $10^{-14}$ | 1.3 x $10^{-6}$ | 8.9 x $10^{-4}$ |

| | | | | | | | | |
|---|---|---|---|---|---|---|---|---|
| ShiftMean_$T_R$ | | | | | | | | |
| TruncateMax_$T_H$ and ShiftMean_$T_H$ | 5.9 x $10^{-13}$ | 1.2 x $10^{-15}$ | 3.7 x $10^{-8}$ | 1.5 x $10^{-5}$ | 2.6 x $10^{-11}$ | 5.2 x $10^{-14}$ | 1.7 x $10^{-6}$ | 6.8 x $10^{-4}$ |

| | Number of illnesses | | | | Probability of ≥1 illness | Number of deaths | | | | Probability of ≥1 death |
|---|---|---|---|---|---|---|---|---|---|---|
| $T_D, T_R, T_H$ | Median | 5% | 95% | 99% | | Median | 5% | 95% | 99% | |
| (baseline) | 0 | 0 | 1,070 | 475,436 | 24.1% | 0 | 0 | 264 | 119,639 | 18.8% |
| -10%_$T_D$ | 0 | 0 | 382 | 246,532 | 20.4% | 0 | 0 | 94 | 61,677 | 15.6% |
| -10%_$T_R$ | 0 | 0 | 446 | 264,869 | 21.4% | 0 | 0 | 111 | 66,469 | 16.5% |
| -10%_$T_H$ | 0 | 0 | 460 | 251,759 | 21.9% | 0 | 0 | 115 | 62,345 | 16.8% |
| TruncateMax_$T_D$ | 0 | 0 | 182 | 129,861 | 17.8% | 0 | 0 | 45 | 31,817 | 13.4% |
| TruncateMax_$T_R$ | 0 | 0 | 198 | 126,767 | 19.1% | 0 | 0 | 49 | 31,604 | 14.5% |
| TruncateMax_$T_H$ | 0 | 0 | 350 | 143,794 | 21.8% | 0 | 0 | 88 | 41,364 | 16.9% |
| ShiftMean_$T_D$ | 0 | 0 | 609 | 364,916 | 21.8% | 0 | 0 | 153 | 35,792 | 16.6% |
| ShiftMean_$T_R$ | 0 | 0 | 294 | 192,170 | 19.8% | 0 | 0 | 72 | 48,555 | 15.2% |
| ShiftMean_$T_H$ | 0 | 0 | 314 | 192,677 | 20.3% | 0 | 0 | 79 | 49,229 | 15.5% |
| TruncateMax_$T_D$ and ShiftMean_$T_D$ | 0 | 0 | 115 | 88,442 | 16.2% | 0 | 0 | 28 | 21,407 | 12.2% |
| TruncateMax_$T_R$ and ShiftMean_$T_R$ | 0 | 0 | 130 | 87,237 | 17.6% | 0 | 0 | 33 | 21,629 | 13.2% |
| TruncateMax_$T_H$ and ShiftMean_$T_H$ | 0 | 0 | 163 | 66,492 | 19.0% | 0 | 0 | 40 | 16,321 | 14.3% |
